# Perivascular space enlargement accelerates with hypertension, white matter hyperintensities, chronic inflammation, and Alzheimer’s disease pathology: evidence from a three-year longitudinal multicentre study

**DOI:** 10.1101/2023.09.25.23296088

**Authors:** Inga Menze, Jose Bernal, Pinar Kaya, Çağla Aki, Malte Pfister, Jonas Geisendörfer, Renat Yakupov, Michael T. Heneka, Frederic Brosseron, Matthias C. Schmid, Wenzel Glanz, Enise I. Incesoy, Michaela Butryn, Ayda Rostamzadeh, Dix Meiberth, Oliver Peters, Lukas Preis, Dominik Lammerding, Daria Gref, Josef Priller, Eike J. Spruth, Slawek Altenstein, Andrea Lohse, Stefan Hetzer, Anja Schneider, Klaus Fliessbach, Okka Kimmich, Ina R. Vogt, Jens Wiltfang, Claudia Bartels, Björn H. Schott, Niels Hansen, Peter Dechent, Katharina Buerger, Daniel Janowitz, Robert Perneczky, Boris-Stephan Rauchmann, Stefan Teipel, Ingo Kilimann, Doreen Goerss, Christoph Laske, Matthias H. Munk, Carolin Sanzenbacher, Petra Hinderer, Klaus Scheffler, Annika Spottke, Nina Roy-Kluth, Falk Lüsebrink, Katja Neumann, Frank Jessen, Stefanie Schreiber, Emrah Düzel, Gabriel Ziegler

**Author notes:** Address for correspondence: Inga Menze, German Centre for Neurodegenerative Diseases (DZNE), Leipziger Str. 44, 39120 Magdeburg, Germany, Mail. Shared first or last authorship (Inga Menze, Jose Bernal, Stefanie Schreiber, Emrah Düzel, Gabriel Ziegler).

## Abstract

**Background:** Perivascular space (PVS) enlargement in ageing and Alzheimer’s disease (AD) and its exacerbators require further investigation.

**Methods:** We studied centrum semiovale (CSO) and basal ganglia (BG) PVS computationally over three to four annual visits in 557 participants of the DZNE multicentre DELCODE cohort. We tested volumetric changes of PVS in relation to ageing, sex, years of education, hypertension, AD diagnosis, and cerebrospinal-fluid-derived Amyloid and Tau positivity and interleukin 6 (IL-6).

**Results:** PVS volumes increased over time. PVS enlargement was associated with baseline white matter hyperintensities. BG-PVS enlargement was related to age and was faster with hypertension. CSO-PVS volumes increased faster with Amyloid and Tau positivity. Higher CSF IL-6 levels predicted PVS volume expansion in both regions and were associated with accelerated PVS enlargement in individuals with Amyloid and Tau positivity.

**Conclusion:** Our work supports the region-specific involvement of white matter hyperintensities, neurotoxic waste accumulation, and inflammation in PVS enlargement.

## 1 Introduction

Perivascular spaces (PVS) are millimetre fluid-filled cavities that surround small perforating cerebral vessels(1,2). PVS can become large enough to be visible in magnetic resonance imaging (MRI)—a condition that is increasingly described in humans in the context of ageing(3,4), arterial hypertension(5,6), structural (e.g. capillary tortuosity) or functional (e.g. blood flow) cerebral small vessel alterations(7,8), and systemic- or neuroinflammation(9–11). These findings collectively suggest that PVS enlargement might indicate a state of dysregulated brain homeostasis(12).

The aetiology of PVS enlargement in humans remains elusive(8). The hypothesised role of PVS in the removal of metabolic and neurotoxic waste products, including Amyloid-β (Aβ) and tau(12–15), key proteins in the pathogenesis of Alzheimer’s disease (AD), proposes that PVS enlargement reflects compromised PVS function and, by extension, potentially impaired glymphatic clearance(2). In line with the clearance hypothesis, PVS burden has been found to be associated with elevated levels of Aβ and tau(16,17), vascular Aβ deposition in cerebral amyloid angiopathy (CAA)(18–20), as well as with clinical diagnosis of mild cognitive impairment (MCI)(21) and AD(22,23). Moreover, a recent study revealed that microglial activation mediates the association between tau and PVS burden in the centrum semiovale (CSO)(24). The finding suggests that tau-induced neuroinflammation might impair clearance, which could lead to a vicious circle presumably comprising blood-brain-barrier permeability alterations and leakage of peripheral immune cells, further promoting neuroinflammation or neurotoxic waste into the PVS. Markers of cerebral small vessel disease (CSVD)(25) and neuroinflammation(26)—which are found consistently associated with the presence of enlarged PVS(24,27)—are also increasingly discussed as a potential aggravating factor in AD pathology(28,29). Taken together this suggests that PVS alterations could potentially link CSVD, neuroinflammation and AD pathology.

While the aforementioned evidence and deductions are compelling, some studies did not find clear associations of AD diagnosis or pathology with PVS burden(30,31), casting doubt on the involvement of PVS alterations in AD and the validity of the clearance hypothesis. Longitudinal studies that could establish specific factors that mechanistically drive PVS enlargement are scarce due to the persistent methodological challenges to quantify PVS reliably and computationally using repeated measures(32). A few prospective studies in healthy ageing suggest that PVS counts(33,34) and volumes(35) increase over time, with their baseline load contributing to both the progression of PVS and of other markers of CSVD, in particular white matter hyperintensities (WMH)(33,35). Other studies nonetheless found only partial or no evidence for longitudinal PVS frequency changes in healthy ageing(36) or in CSVD(37).

Taken together, these findings point to the relevance of further research into conditions that contribute to the presence and dynamical changes of PVS and their potential interactions, particularly in the context of a complex multifactorial neurodegenerative disease like AD. Leveraging on the largest longitudinal multi-centre study of PVS in humans to date (557 subjects along AD syndromal cognitive stages; 4 annual time points, 2228 multimodal structural MRI scans), we here focus on quantifying PVS enlargement and change dynamics using a multimodal segmentation approach and longitudinal modelling. Our aim, on the one hand, is to characterise dynamical changes of PVS in healthy ageing and, on the other hand, to determine individual effects of WMH, hypertension, biomarkers of AD pathology, and inflammation.

## 2 Methods and Materials

### 2.1 Study design and participants

We used baseline, 12-, 24-, and 36-month follow-up data from DELCODE (DZNE Longitudinal Cognitive Impairment and Dementia Study(38); **Supplementary Figure S1**)—an observational multicentre study from the German Centre for Neurodegenerative Diseases (DZNE) that uses multimodal assessment of preclinical, prodromal, and clinical stages of AD, with a particular focus on subjective cognitive decline. We focused on non-complaining, asymptomatic healthy controls with normal cognition (NC), cognitively normal first-degree siblings of AD patients (ADR), participants with subjective cognitive decline (SCD), mild cognitive impairment (MCI), and AD patients. We additionally restricted our sample to participants that attended at least three MRI scanning sessions to reliably estimate PVS rates of change (n = 557; 2228 sets of T1w and FLAIR).

All participants entered DELCODE based on either their clinical diagnosis derived from the clinical workup or their identification as asymptomatic control subjects or ADR. Before joining DELCODE, participants received an extensive assessment at the local study site, which included medical history, psychiatric and neurological examination, neuropsychological testing, assessment of blood and cerebrospinal fluid (CSF) markers of neurodegeneration and inflammation, and routine MRI in accordance with local standards. All memory clinics used the Consortium to Establish a Registry for AD (CERAD-plus) neuropsychological test battery to assess cognitive function.

SCD, amnestic MCI, and AD diagnoses followed the existing research criteria(38). Participants with SCD reported subjective cognitive decline or expressed memory concerns to the physician of each memory centre but had a cognitive performance above -1.5 standard deviations (SD) below the age, sex, and education-adjusted normal performance on all subtests of the CERAD-plus test battery. Participants with amnestic MCI obtained an age-, sex-, and education-adjusted performance below -1.5 SD on the delayed recall trial of the CERAD-plus word-list episodic memory tests. Subjects with AD fulfilled NINDCS/ADRDA criteria and had a CERAD-plus score of below -1.5 SD, an extended MMSE score between 18-26, and a CDR rating greater or equal to one. ADR were included in DELCODE if their performance was within 1.5 SD in the CERAD-plus test battery.

Recruitment of the NC and ADR group took place through local newspaper advertisement and screening for subjective cognitive impairment via telephone. Participants in the NC group were subjectively unimpaired and had objective cognitive performance similar to that of the SCD group. Very subtle cognitive impairment, which did not cause concerns and was considered normal for the participant’s age, was not an exclusion criterion for NC.

Additional inclusion criteria for all groups were age ≥ 60 years, fluent German language skills, capacity to provide informed consent, and presence of a study partner. The main exclusion criteria for all groups were conditions clearly interfering with participation in the study or the study procedures, including significant sensory impairment. The following medical conditions were considered exclusion criteria: current major depressive episode, major psychiatric disorders either at baseline or in the past (e.g., psychotic disorder, bipolar disorder, substance abuse), neurodegenerative disorders other than AD, vascular dementia, history of stroke with residual clinical symptoms, history of malignant diseases, severe or unstable medical conditions, and clinically significant abnormalities in vitamin B12. Prohibited drugs included chronic use of psychoactive compounds with sedative or anticholinergic effects, use of anti-dementia agents in SCD, amnestic MCI, and NC subjects, and investigational drugs for the treatment of dementia or cognitive impairment one month before entry and throughout the duration of the study.

All participants gave written informed consent in accordance with the Declaration of Helsinki prior joining the study. DELCODE is retrospectively registered at the German Clinical Trials Register (DRKS00007966, 04/05/2015). Ethics committees of the medical faculties of all participating sites, Berlin (Charité, University Medicine), Bonn, Cologne, Göttingen, Magdeburg, Munich (Ludwig-Maximilians-University), Rostock, and Tübingen, gave ethical approval for this work. The ethics committee of the medical faculty of the University of Bonn led and coordinated the process.

#### 2.1.1.1 Classification of groups

Throughout the manuscript, we refer to four specific subsamples: a *presumably healthy* subsample (NC and SCD, i.e., those without objective cognitive impairment), a subsample of *possible genetic predisposition* (ADR only), and two *clinical symptomatic groups* (MCI and AD).

#### 2.1.2 Hypertension

We categorised subjects into normotensive and hypertensive according to their ICD-10 diagnosis, as described in(39). This information was available for 555 subjects (99.64%). As most subjects with hypertension (n = 298, 53.69%) had been prescribed antihypertensive medication (n = 286, 95.97%), we refer to this group as treated hypertensive subjects.

#### 2.1.3 CSF biomarker assessment

CSF biomarker samples were obtained through lumbar puncture by trained study assistants, centrifuged, aliquoted, and stored at -80°C for retests, and analysed with commercially available kits (V-PLEX Aβ Peptide Panel 1 (6E10) Kit (K15200E); Innotest Phospho Tau(181P), 81581, Fujirebio Germany GmbH, Hannover, Germany). CSF biomarker data were available for 268 participants.

##### 2.1.3.1 Alzheimer’s disease biomarker profiles

We determined each participant’s Amyloid- and Tau-positivity based on the Aβ42/40 ratio and phosphorylated tau181 (pTau181) levels in CSF. DELCODE-specific cut-offs for biomarker positivity were established from baseline data via Gaussian mixture modelling (Amyloid-negative: Aβ42/40 > 0.08; Amyloid-positive: Aβ42/40 ≤ 0.08; Tau-negative: pTau181< 73.65; Tau-positive: pTau181 ≥ 73.65)(40).

We classified individuals on the AD continuum based on their Amyloid-(A) and Tau-positivity status (T) into: A-T-, A-T+, A+T-, and A+T+(28) (n = 167, 6, 56, 39, respectively). In analyses investigating PVS enlargement in relation to these AD biomarker profiles, we excluded the A-T+ group since it had a small sample size and it is not considered to reflect AD pathological change(28).

##### 2.1.3.2 Inflammation

CSF biomarker samples also enabled measuring CSF interleukin-6 (IL-6) concentrations. These were conducted using immunoassays (Quanterix Simoa Singleplex Assay (HD-1)) optimised for detection range of the respective markers using technical duplicates, coefficient of variance < 20%, and inter-run-normalisation by internal control samples(41). CSF IL-6 levels were available in a subsample of n = 184 subjects (n = 65, 58, 17, 32, 12 for NC, SCD, ADR, MCI, AD respectively).

#### 2.1.4 Structural magnetic resonance imaging

MRI acquisition took place at nine DZNE sites equipped with 3T Siemens MR scanners. In the present work, we leveraged T1w MPRAGE (full head coverage; 3D acquisition, GRAPPA factor 2, 1 mm^3^ isotropic, 256 × 256 px, 192 sagittal slices, TR/TE/TI 2500/4.33/1100 ms, FA 7°), T2w partial head coverage (0.5 × 0.5 × 1.5 mm, 384 × 384 px, 64 quasi-coronal slices perpendicular to hippocampal long axis, TR/TE 3500/353 ms), T2w turbo spin-echo (full head coverage; 0.8 × 0.8 × 2 mm, 240 × 320 px, 72 axial slices, TR/TE 6500/79 ms) and T2w FLAIR (full head coverage; 1 mm^3^ isotropic, 256 × 256 px, 192 sagittal slices, TR/TE/TI 5000/394/1800 ms) images. The DZNE imaging network oversaw operating procedures and quality assurance and assessment (iNET, Magdeburg)(38).

### 2.2 Segmentation and quantification

#### 2.2.1 Regions of interest

We generated whole-brain parcellations and white matter segmentations for each subject and for each visit using FreeSurfer (v7.1). We used the resulting segmentation maps to define basal ganglia (BG) and CSO regions, as per Potter’s scale(42).

#### 2.2.2 Hippocampal volume

We estimated hippocampus volumes across time using FreeSurfer 7.1 longitudinal processing(43).

#### 2.2.3 White matter hyperintensities volume

We estimated fractional WMH volumes ([WMH volume / ROI volume] * 100) using the Lesion Prediction Algorithm in the Lesion Segmentation Toolbox(44) and FLAIR imaging data.

#### 2.2.4 Multi-modal PVS segmentation

We segmented PVS computationally using a multimodal approach in order to alleviate quantification issues that the presence of WMH often pose(45,46) (Supplementary information on “computational PVS segmentation” including **Figures S2** and **S3** provides algorithms and methods). We particularly took inspiration from works examining T1w and FLAIR imaging sequences to improve distinguishing between these two neuroradiological features(21,47), in line with the STRIVE-2 recommendations(48). The process consisted of denoising all input images with a non-local means filtering technique(49), enhancing tubular structures with the Frangi filter(50), fusing T1w and FLAIR information to better distinguish PVS from WMH, and binarising with white matter (WM) and BG specific thresholds. We report fractional PVS volumes ([PVS volume / ROI volume] * 100), which indicate how many percent of the region of interest (ROI) corresponds to PVS.

##### Segmentation parameter tuning

We optimised PVS segmentation thresholds that maximised the correlation between qualitative and computational estimates of PVS burden, as in(51). Two neurology residents (MP and JG) blinded to clinical data rated PVS severity in CSO and BG independently on T2w images with partial head coverage using the Potter’s rating procedure(42).

##### Clinical validation

In 30 randomly drawn subjects (11 females; mean age 70 (*SD* 4.35) years; n = 1, 3, 21, 4, 1 for NC, ADR, SCD, MCI and AD, respectively) a third independent neurology resident (PK), blinded to clinical data, counted PVS in the T2w full head coverage axial slice with the highest burden separately for CSO and BG and we compared them to our estimated computational counts in the same slice and estimated fractional volumes. We used correlations, Lin’s concordance correlation coefficient (CCC), and Bland-Altman plots for this purpose.

We also assessed stability of computational PVS segmentation over repeated measures by correlating fractional PVS volumes—controlled for age, sex, education and clinical diagnosis—across time points.

### 2.3 Hypotheses and statistical analyses

#### 2.3.1 Adjusting baseline PVS volumes

We corrected baseline fractional PVS volumes for linear and quadratic age effects, sex, and years of education via residualisation.

#### 2.3.2 Estimating PVS rates of change

We estimated *rates of change* in fractional CSO- and BG-PVS volumes using linear mixed effect (LME) modelling. All LME models included correlated random intercepts and random slopes. We followed the recommendations of Guillaume et al.(52), i.e. we incorporated a longitudinal ’visit’—or here *time*—effect (centred intra-subject age) and a cross-sectional ’age’ effect into our models. We controlled for linear and quadratic age effects, sex and years of education. We visually screened residual plots and normality via Q-Q plots to check linearity and homoscedasticity. We excluded outliers (standard residual greater than 3 SD) and influential data points exceeding a cut-off value suggested by Van der Meer and colleagues(53) (Cook’s distance > 4/n). We reported effect size estimates using beta coefficients, 95% confidence intervals and an alpha level of *p* < 0.05. We extracted subject-specific slopes from the fitted LME models, which denote the individual rate of change in fractional PVS volumes over time. Hence, subject-specific rates of change encompassed the combination of fixed and random effects.

We additionally reported annual rates of change of fractional and absolute CSO- and BG-PVS volumes for each clinical group (in % / year and mm³ / year, respectively) in order to provide a rough yet interpretable estimate of change. For each clinical group, we computed fractional PVS volumes at each time point, corrected these estimates for age, and estimated the annual rates of change with a linear model. We then converted these rates to absolute change of PVS volumes in mm³ / year by multiplying them by the average ROI volume (in mm³) within the diagnostic group.

#### 2.3.3 Effects of further contributing factors

##### Ageing

PVS visibility—and thus volumes—increase with ageing(7,33,35,36). In spite of this, recent cross-sectional studies across the lifespan also suggest that such a process can be of second-order (polynomial) nature(3,4), making both acceleration and saturation effects conceivable. Though elusive, the scale at which enlargement occurs over short time periods, limitations in imaging resolution (e.g. partial volume effects), and methodological confounds caused by the presence or formation of WMH around PVS(54) may help to explain this situation. Accordingly, we tested for linear and quadratic effects of age on PVS measurements in the presumably healthy subsample, to assess supposedly pathology-free ageing processes. Using multiple linear regression, we tested for said associations with baseline fractional PVS volumes cross-sectionally. As described above, in LME models we accordingly evaluated associations of change in fractional PVS volumes over time with linear and quadratic age effects.

##### WMH

Numerous cross-sectional studies support the positive association between PVS and WMH(2) and a few even provide evidence of WMH developing in the proximity of PVS(54). We used correlational analysis to determine whether baseline regional WMH volumes explained part of the variance in regional PVS rates of change in the presumably healthy subsample, after adjusting for age, sex, and years of education.

##### Hypertension

Lenticulostriate arteries in the BG transmit higher blood pressure than those in CSO, rendering them more vulnerable to the repercussions of arterial hypertension(55). We therefore expected that subjects with a history of hypertension had increased fractional PVS volumes at baseline and steeper volume increases than those without one. We tested these effects by comparing baseline fractional PVS volumes and PVS rates of change in treated hypertensive versus normotensive cases of our presumably healthy subsample using the Mann-Whitney-U Test.

We also tested for interactive effects of the risk factors hypertension (normotensive vs. treated hypertensive) and clinical diagnosis (presumably healthy vs. ADR vs. clinical symptomatic groups) on PVS volumes at baseline and their rates of change via 2×4 ANOVAs. We identified outliers (Q3 + 1.5×IQR or below Q1 - 1.5×IQR of group median) and removed them prior to ANOVA.

##### AD pathology

Since malfunctioning of brain clearance pathways—in which PVS are thought to play a part(12–15)—may result in pTau and Aβ accumulation as well as enlargement of PVS around perforating cortical vessels(17,19,20), we anticipated CSO-PVS rates of change to be higher in A+T- and A+T+ versus A-T-, and, consequently, fractional volumes and trajectories to be higher in AD versus NC (16,17,22,23). We note, however, that vicious cycles are likely, i.e., neurotoxic waste accumulation can promote clearance pathway failure and vice versa. We used Kruskal-Wallis-tests and Bonferroni-corrected Dunn-tests for pairwise comparisons to analyse differences of rates of change across diagnostic groups and AD biomarker profiles. We used the same approach to additionally test for baseline differences across diagnostic groups.

We also investigated whether baseline PVS volume and PVS rates of change related to hippocampal volume and hippocampal atrophy rates, respectively. We estimated total hippocampal volume by aggregating volumes across hemispheres and correcting for total intracranial volume (TIV) via residualisation. We obtained hippocampal rates of change using a similar LME model as for PVS (*see 2.3.2 Estimating PVS rates of change*), in which we adjusted for linear and quadratic age effects, sex, and years of education. The LME model used to extract hippocampal volume rates of change can be found in Supplementary **Table S4**.

##### Inflammation

Cross-sectional imaging studies support associations of PVS with inflammatory markers(9,10), with histopathological studies even suggesting PVS are sites of immune cell infiltration and cytokine accumulation (*for reviews see*(2,56)). Here, we focused particularly on CSF interleukin-6 (IL-6)—a pro-inflammatory cytokine released by peripheral immune cells(29) and moreover associated with microglia activation and their modulation(26)— since it has been found linked to chronic neuroinflammation across multiple age-related pathologies(57), including AD(26), and to relate to qualitative PVS severity estimates, esp. in the BG(11). Because IL-6 appears to be released in relation to Aβ accumulation (*for review see*(29)), and relates to aggravation of markers of CSVD(24,27), we also tested for interactive effects of CSF IL-6 levels and AD biomarker status on PVS rates of change. For this purpose, we first evaluated associations between PVS rates of change and CSF IL-6 by extending our LME models by an interaction of time and CSF IL-6. Second, we fitted models with additional fixed effects of AD biomarker profile, CSF IL-6 levels, a two-way interaction of time and CSF IL-6 levels, as well as a three-way interaction of AD biomarker profile, CSF IL-6 levels and time. The latter was of main interest to assess interaction effects between AD biomarker profile and CSF IL-6 level on PVS rates of change. We focused on differences between A-T- and A+T+. We used A+T+ subjects as our reference group.

#### 2.3.4 Healthy ageing versus AD effects

With the aim of disentangling the contributions of ageing alone but also in conjunction with AD pathology, we conducted two separate statistical analyses: one for the presumably healthy ageing subsample (NC, SCD) and one on the whole cohort, which further encompassed symptomatic stages (MCI, AD) and ADR.

#### 2.3.5 Data transformation

We transformed fractional PVS volumes using the Box-Cox transformation (model with intercept only) to deal with non-normality and skewness. We also z-scored years of education and age, and log-transformed fractional WMH volumes and IL-6 concentration.

#### 2.3.6 Software

We analysed data with *R* (v4.0.2) using RStudio (v1.3.1073)(58). We modelled and diagnosed linear mixed effect models (LME) with *lme4*, *lmerTest*, *influence.ME* and *psych*. Correlations and group differences (Mann-Whitney U-tests, Kruskal Wallis tests, ANOVAs) were conducted with *rstatix.* We created figures with *ggplot2*, *ggpubr*, and *ITK-SNAP* (59).

## 3 Results

### 3.1 Clinical validation and reliability of repeated measures of PVS

We assessed whether computational and clinical PVS assessments aligned after optimising thresholding parameters for both BG (1.0309×10^-4^) and WM (2.7284×10^-5^). We obtained moderate polyserial correlations between fractional PVS volumes and visual scores (WM: 0.54; BG: 0.51; **Figure 1A, B**).

**Figure 1.**
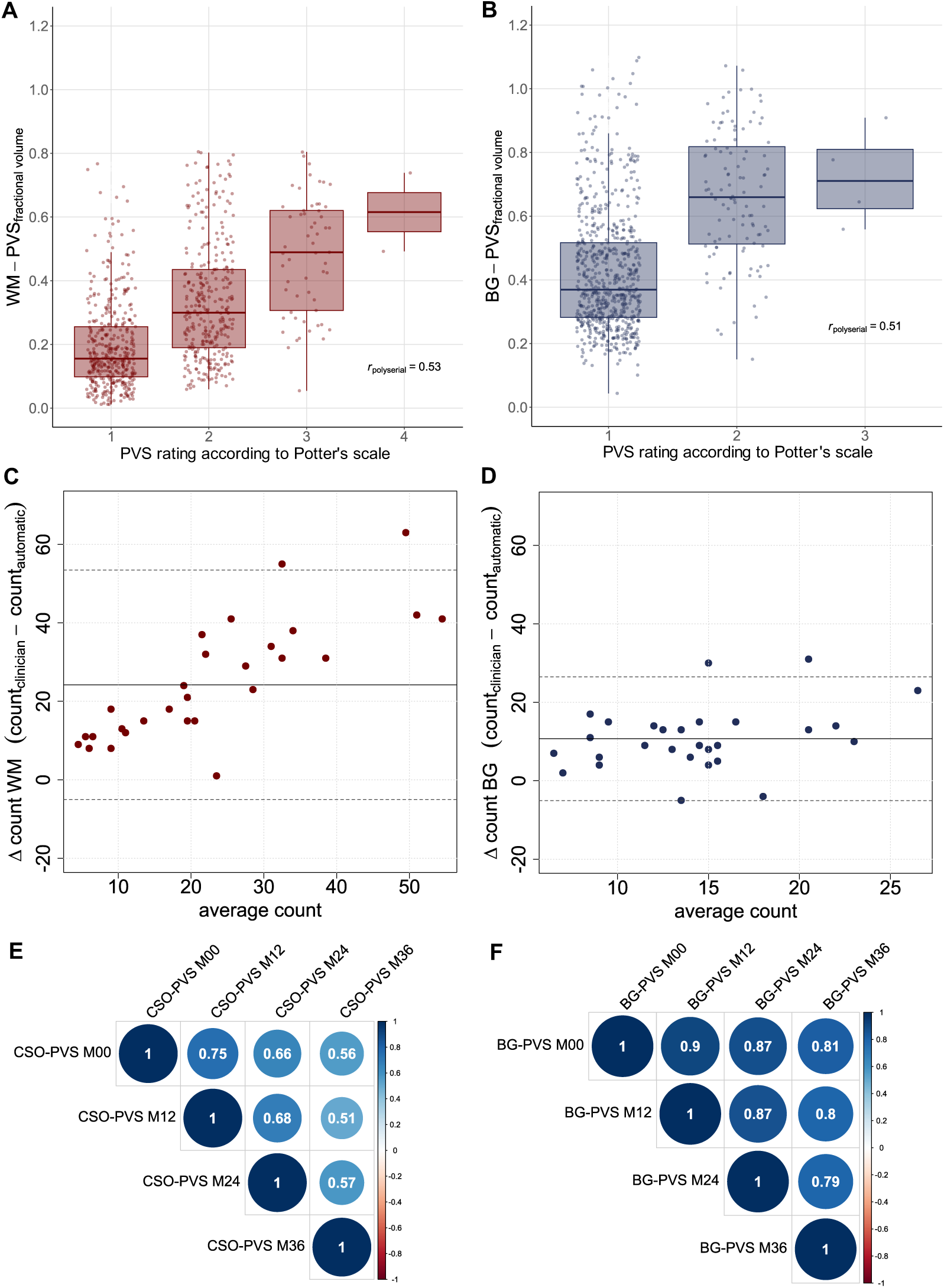
Clinical validation and reliability of repeated measures. (**A-B**) Moderate polyserial correlations between fractional PVS volumes and visual scores. (**C-D**) Bland-Altman plot comparing manual and computational PVS counts in the axial slice with the highest PVS burden in white matter and basal ganglia in subset of 30 random subjects. Solid lines depict the mean difference and dotted lines depict the corresponding 95%-confidence intervals. (**E-F**) High Pearson’s correlations of fractional CSO- and BG-PVS volumes across four annual time points suggest measurement stability. We corrected fractional PVS volumes for age, years of education, sex and clinical diagnosis and used the Box-Cox transformation to deal with skewness.

In the subset of 30 randomly-selected subjects, computational and manual counts in the slice with the highest PVS burden correlated strongly in WM (*ρ_spearman_* = 0.75, *p* < 0.001) and moderately in BG (*ρ_spearman_* = 0.21, *p* = 0.264). Manual counts and computational fractional volumes exhibited a similar relationship (WM: *ρ_spearman_* = 0.70, *p* < 0.001; BG: *ρ_spearman_* = 0.36, *p* = 0.052). We noticed an underestimation of PVS counts by our computational approach (WM: *difference*= 24.20, *SD* 14.93; BG: *difference* = 10.72, *SD* 8.04), which resulted in low Lin’s concordance correlation coefficients (WM: *CCC* = 0.25, [*95%-CI* 0.12, 0.37]; BG: *CCC* = 0.08 [*95%-CI* -0.07, 0.22]). In addition, the Bland-Altman plot suggested a proportional error: the more PVS there were, the fewer PVS the method detected (**Figure 1C, D**). Given that PVS sensitivity is modality specific, said underestimation is to be expected.

Correlation of computational PVS segmentation across repeated measures was high for both CSO (**Figure 1E**) and BG (**Figure 1F**) suggesting a moderate to high reliability.

### 3.2 Descriptive statistics and sample characteristics

We studied PVS in 557 DELCODE participants (**Figure S1;** sample characteristics in **Table 1**). Fractional PVS volumes in BG and CSO increased by at least 0.022 % according to a linear approximation of the annual change (**Table 1**). In the CSO, annual changes in absolute PVS volumes were the highest for AD (AD ≈ 17.05 mm³ / year vs NC ≈ 7.04 mm³ / year). In the BG, annual change rates in fractional PVS volumes were similar across groups (≈ 0.02 %), regardless of atrophy levels.

**Table 1.** Descriptive statistics for fractional CSO-PVS and BG-PVS volumes for four annual visits, stratified by clinical groups. We reported values on subjects who had at least three scans available (n=557). We removed extreme values in fractional volumes within the whole sample for each time point to lessen the influence of outliers and corrected estimates for age at the time of the scan.

| Clinical group | N | Baseline demographics |  |  | Baseline |  | 12-month follow-up |  | 24-month follow-up |  | 36-month follow-up |  | Annual change* of PVS volumes mm<sup>3</sup> / year<sup>†</sup><br>(%fractional volumes / year) |  |
| --- | --- | --- | --- | --- | --- | --- | --- | --- | --- | --- | --- | --- | --- | --- |
|  |  | Females (%) | Age (years) | Education (years) | Fractional volume (%) |  | Fractional volume (%) |  | Fractional volume (%) |  | Fractional volume (%) |  | CSO-PVS | BG-PVS |
|  |  |  |  |  | CSO-PVS | BG-PVS | CSO-PVS | BG-PVS | CSO-PVS | BG-PVS | CSO-PVS | BG-PVS |  |  |
| NC | 166 | 54.82 | 69.2±0.38 | 14.9±0.22 | 0.14±0.01 | 0.46±0.02 | 0.15±0.01 | 0.48±0.02 | 0.16±0.01 | 0.48±0.02 | 0.21±0.02 | 0.55±0.02 | 7.04<br>(0.022) | 18.67<br>(0.027) |
| ADR | 51 | 56.86 | 65.6±0.60 | 14.4±0.39 | 0.21±0.02 | 0.49±0.04 | 0.23±0.02 | 0.50±0.04 | 0.24±0.03 | 0.52±0.04 | 0.31±0.04 | 0.57±0.05 | 10.59<br>(0.031) | 18.10<br>(0.026) |
| SCD | 238 | 48.74 | 70.9±0.39 | 15.0±0.19 | 0.14±0.01 | 0.49±0.02 | 0.16±0.01 | 0.52±0.02 | 0.20±0.01 | 0.54±0.02 | 0.21±0.02 | 0.56±0.02 | 7.92<br>(0.025) | 16.03<br>(0.023) |
| MCI | 71 | 40.85 | 72.5±0.69 | 14.0±0.37 | 0.16±0.02 | 0.53±0.03 | 0.17±0.02 | 0.53±0.03 | 0.18±0.02 | 0.58±0.03 | 0.26±0.04 | 0.59±0.04 | 8.80<br>(0.031) | 15.34<br>(0.023) |
| AD | 31 | 54.84 | 74.3±1.12 | 12.9±0.50 | 0.16±0.03 | 0.58±0.04 | 0.22±0.03 | 0.55±0.05 | 0.26±0.04 | 0.62±0.05 | 0.38±0.07 | 0.63±0.08 | 17.05<br>(0.070) | 13.82<br>(0.022) |

Annotations. Mean ± standard error. \*Annual change in fractional volume estimated under linear assumptions and based on age-corrected untransformed fractional PVS volumes per diagnostic group. †Absolute change of PVS volumes in mm<sup>3</sup>/year per diagnostic group was calculated based on the annual change of fractional PVS volumes per year and the mean ROI volume within diagnostic group in mm<sup>3</sup>.

### 3.3 Factors contributing to PVS dynamics

#### 3.3.1 Presumably healthy ageing (NC and SCD only)

Cross-sectional multiple linear regression revealed both a linear (*B* = 0.16 [*95%-CI* 0.09, 0.23], *p* < 0.001) and a negative quadratic age effect (*B* = -0.10 [*95%-CI* -0.17, -0.02], *p* = 0.011) contributing to fractional BG-PVS volumes (**Figure 2C**).

**Figure 2.**
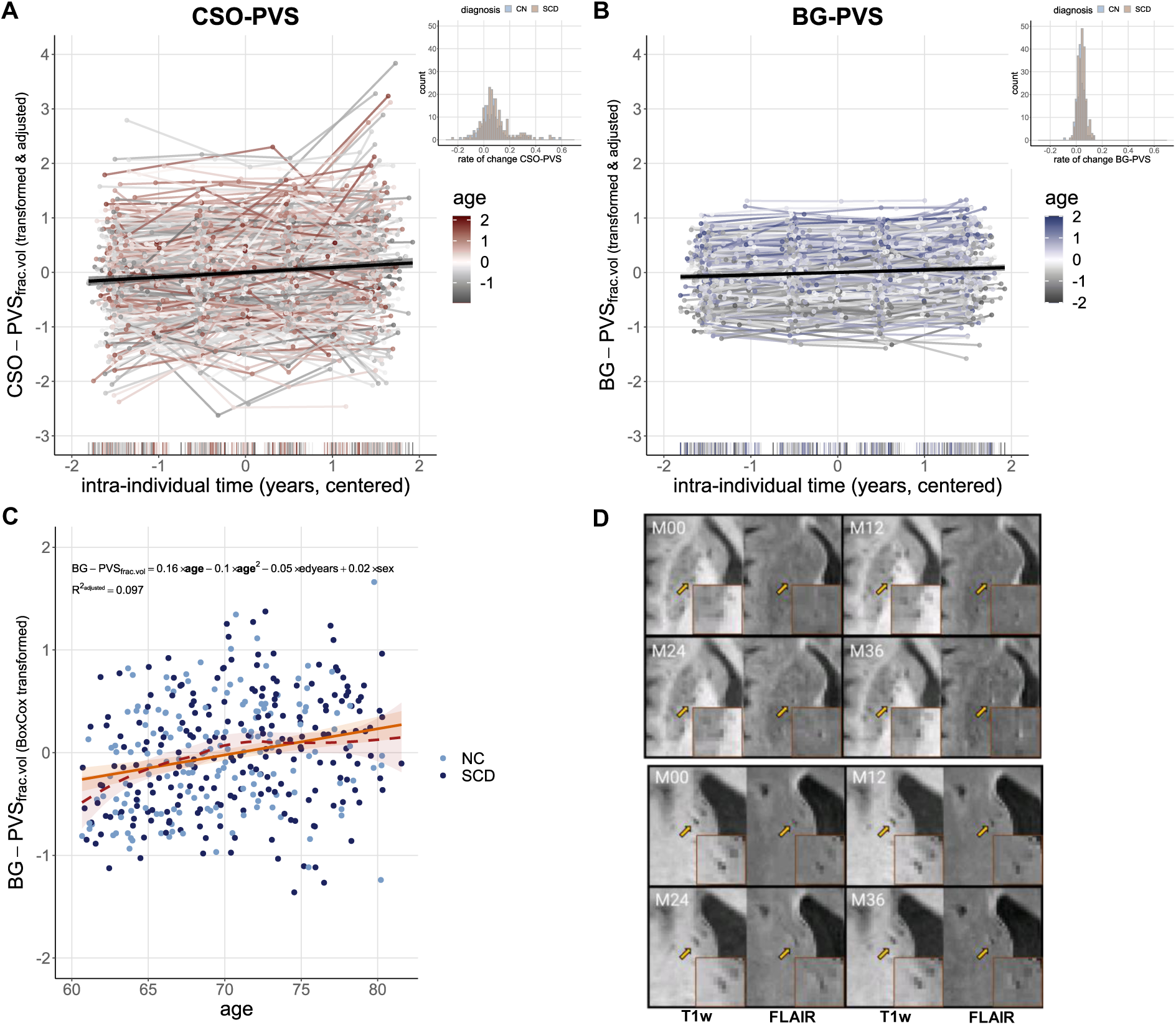
PVS enlargement over follow-ups and with advancing age in presumably healthy ageing (NC and SCD). (**A**-**B**) Fractional CSO- and BG-PVS volumes increased over time (CSO: *B*=0.08 [*95%-CI* 0.06-0.11], *p*<0.001; BG: *B* = 0.04 [*95%-CI* 0.03, 0.05]), *p* < 0.001). Histograms show respective distribution of rates of change in SCD and NC. For easier visual comparison of individual trajectories of PVS volumes across BG and CSO, BoxCox transformed PVS volumes were additionally z-transformed. Plotted fractional PVS volumes were furthermore adjusted for effects of sex and education. (**C**) Cross-sectional associations of linear (orange solid line) and negative quadratic (red dotted line) age effects with fractional BG-PVS volumes. (**D**) Within a three-year period, WMH may emerge in the vicinity of PVS. Example images of a SCD patient from two different axial slices.

Longitudinal analysis via LME (**Table 2**) confirmed linear (*B* = 0.19 [*95%-CI* 0.14, 0.24], *p* < 0.001) and negative quadratic associations of age (*B* = -0.07 [*95%-CI* -0.12, -0.02], *p* = 0.012) with fractional BG-PVS volumes—the latter hinting at potential saturation effects in this specific brain region. Importantly, our longitudinal analysis revealed that fractional BG- and CSO-PVS volumes increased over time (CSO: *B* = 0.08 [*95%-CI* 0.06, 0.11], *p* < 0.001; BG: *B* = 0.04 [*95%-CI* 0.03, 0.05], *p* < 0.001; **Figure 2A, B**). Moreover, female sex was by tendency negatively associated with fractional CSO-PVS volumes

**Table 2.**
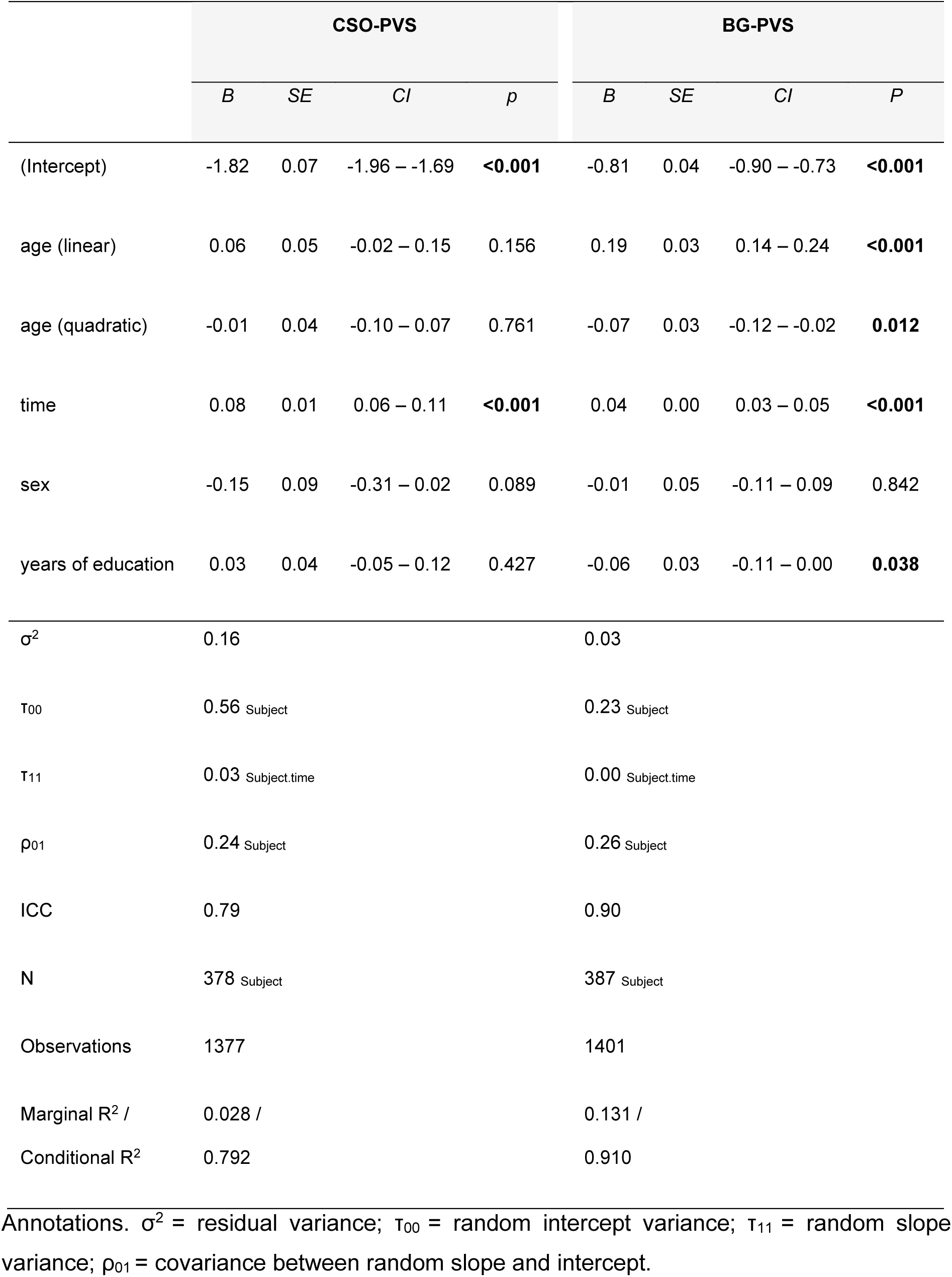
Linear mixed effect modelling for CSO-PVS and BG-PVS in presumably healthy subsample, showing different trajectories over time, effects of age, sex and years of education. Models with correlated slope and random intercept: PVS ∼ age + age² + time + sex + years of education + (1+ time | subject).

(*B* = -0.15 [*95%-CI* -0.31, 0.02], *p* = 0.089); such sex differences were nonetheless not evident for BG-PVS. BG-PVS volumes were lower in subjects with more years of education (*B* = -0.06 [*95%-CI* –0.11, 0.00], *p* = 0.038).

Furthermore, we observed higher interindividual variability in random intercepts (0.56) and slopes (0.03) for CSO-PVS as compared to BG-PVS (random intercept = 0.23; random slope = 0.00; *see histograms in* **Figure 2A, B**).

##### 3.3.1.1 Baseline PVS and rates of change across regions

Correlations between baseline estimates (*ρ* = 0.14, *p_FDR_* = 0.001) and rates of change (*ρ* = 0.24, *p_FDR_* < 0.001) of fractional CSO-and BG-PVS volumes in the presumably healthy subsample support a shared underlying mechanism.

##### 3.3.1.2 White matter hyperintensities

Baseline WMH and fractional PVS volumes were associated with one another across regions (all *ρ* > 0.19, *p_FDR_* < 0.001). Moreover, subjects with the highest WMH volumes at baseline had higher PVS rates of change (CSO: *ρ* = 0.09, *p_uncorrected_* = 0.044, *p_FDR_* = 0.067; BG: *ρ* = 0.16, *p_uncorrected_* = 0.002, *p_FDR_* = 0.003). Visual inspections provided evidence of WMH forming around PVS (**Figure 1D**).

##### 3.3.1.3 Hypertension

Baseline fractional PVS volumes (CSO: *W* =19931, *p* = 0.647, *r* = 0.023; BG: *W* =19567, *p* = 0.891, *r* = 0.007) and PVS rates of change in CSO (*W* =17966, *p* = 0.719, *r* = 0.019) did not differ between treated hypertensive and normotensive subjects in the presumably healthy subsample. Fractional BG-PVS rates of change were nonetheless higher in treated hypertensive compared to normotensive subjects (BG: *W* = 16036, *p* = 0.028, *r* = 0.112).

##### 3.3.1.4 Inflammation

Our LME models additionally accounting for main effects of CSF IL-6 and its interaction by time revealed a non-significant interaction of time by CSF IL-6 levels for CSO-PVS (*B* = 0.07 [*95%-CI* -0.02, 0.16], *p* = 0.118; **Supplementary table S5**), and a trend-wise interaction for BG-PVS (*B* = 0.03 [*95%-CI* -0.01, 0.07], *p* = 0.094; **Supplementary table S5**), indicating limited evidence for accelerated increase of PVS volumes over measurements with higher baseline CSF IL-6 levels in our presumably healthy subsample.

### 3.4 Alzheimer’s disease pathology

When probing the association of fractional PVS volumes with AD diagnosis and with pathology, we observed regional differences. LME models for the full sample, which were used to derive subject-specific slopes for group comparisons can be found in **Supplementary table S6**. In the CSO, despite baseline fractional volumes showing only trends towards differences across clinical groups (*Χ²*(4) = 9.03, *p* = 0.060, *η²* = 0.02), participants had higher CSO-PVS rates of change if they were further along the AD syndromal cognitive stages (*Χ²*(4) = 14.50, *p* = 0.006, *η²* = 0.02). Indeed, CSO-PVS volumes increased at a faster rate in AD versus NC (*p_Bonferroni_* = 0.004; see also **Table 1** for approximation of annual change of PVS volumes and **Figure 3A** for distribution of CSO-PVS rates of change across clinical groups). The AD biomarker profiles explained, in part, these group differences (*Χ²*(2) = 7.29, *p* = 0.026, *η²* = 0.02): CSO-PVS rates of change were lower in A-T-as compared to A+T+ (*p_Bonferroni_* = 0.021; **Figure 3C, D**). Hippocampal volume at baseline or hippocampal rates of change did not significantly correlate with CSO-PVS at baseline or rates of change.

**Figure 3.**
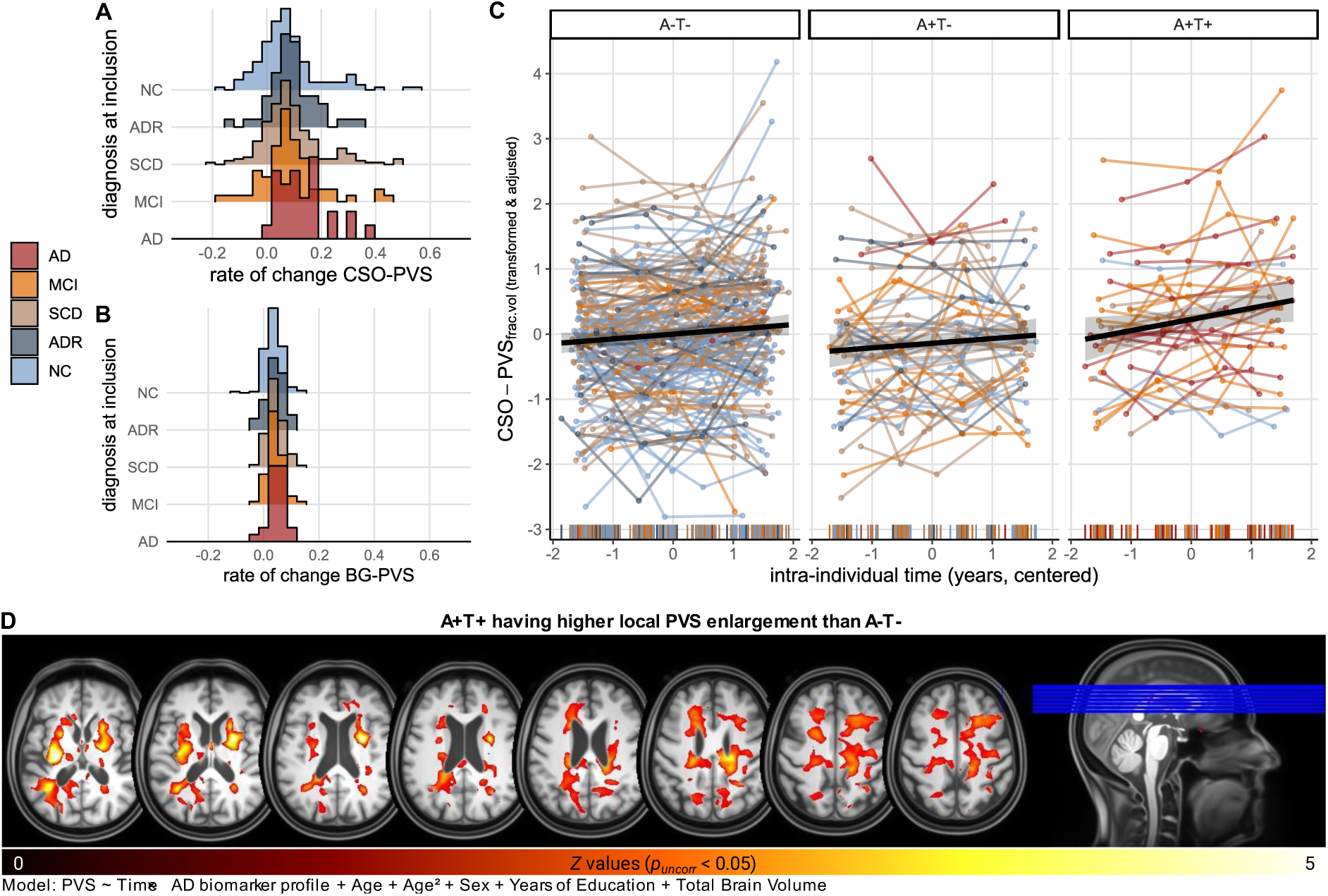
CSO-PVS enlargement is more pronounced in individuals with AD diagnosis and pathology. (**A**-**B**) Histogram ridgeline plots of distribution of rates of change across clinical groups shows higher variance and higher rates of change further along the AD syndromal cognitive stages for CSO-PVS as compared to BG-PVS. (**C**) Increase of fractional CSO-PVS is dependent on AD biomarker profile. Colour of individual trajectories corresponds to clinical diagnosis at time of entry to DELCODE study. Subjects with A+T+ status show higher rates of change as compared to A-T-. Plotted fractional PVS volumes (BoxCox transformed) were additionally *z*-transformed and adjusted for effects of sex, education and linear as well as quadratic age. (**D**) Contrast image highlighting regions where PVS enlargement was more evident in A+T+ vs. A-T- (*p_uncorr_* < 0.05; further information on Supplementary Material on “computational PVS segmentation: 4. Spatial processing for marginal modelling”).

In the BG, on the other hand, we did not find sufficient evidence for a link between BG-PVS and the AD pathology: baseline fractional volumes (*Χ²*(4) = 3.25, *p* = 0.517, *η²* = 0.005), and rates of change (*Χ²*(4) = 3.56, *p* = 0.469, *η²* = 0.006) did not differ between groups (see also **Figure 3B** for distribution of BG-PVS rates of change across clinical groups). Moreover, AD biomarker profiles did not explain variance in BG-PVS rates of change (*Χ²*(2) = 3.01, *p* = 0.222, *η²* = 0.004).

#### 3.4.1 Interaction between AD and hypertension

We evaluated possible interaction effects of clinical diagnosis and hypertension on PVS volumes at baseline and their rates of change via ANOVAs. We did not find significant interactions of hypertension and clinical diagnosis for baseline PVS volumes (CSO: *F*(3,520) = 1.72, *p* = 0.162, *η²* = 0.010, BG: *F*(3,537) = 1.24, *p* = 0.294, *η²* = 0.007). While rates of change in CSO-PVS were unaffected by such interactions (*F*(3,464) = 0.260, *p* = 0.854, *η²* = 0.002), we detected a marginally significant interaction effect for rates of change in BG-PVS (*F*(3,508) = 2.47, *p* = 0.061, *η²* = 0.014). This finding confirmed the influence of hypertension on BG-PVS rates of change in the presumably healthy sample (NC, SCD, *p* = 0.060), while it did not exhibit a modulatory effect on BG-PVS rates of change within clinical groups.

#### 3.4.2 Inflammation

##### 3.4.2.1 Interaction between inflammation and AD biomarkers

Not distinguishing by AD biomarker profile, we observed accelerated increase of PVS volumes over measurements with higher baseline CSF IL-6 levels in the whole sample (**Figure 4A, B**; see also **Supplementary tables S7, S8** for time × CSF IL-6 interactions on whole sample). To evaluate the interaction effect of interest between AD biomarker profile and CSF IL-6 level on PVS rates of change, we extended the previous LME models by a three-way interaction of AD biomarker profile, CSF IL-6 levels and time. In A+T+, the increase of CSO- and BG-PVS volumes over measurements was higher with higher baseline CSF IL-6 levels (CSO: time x CSF IL-6: *B* = 0.12 [*95%-CI* 0.02, 0.21], *p* = 0.016, **Figure 4C**; BG: time x CSF IL-6: *B* = 0.05 [*95%-CI* 0.01, 0.09], *p* = 0.009, **Figure 4D**). For CSO-PVS, this modulation appeared lower in A-T-subjects (time x CSF IL-6 x A-T-: *B* = -0.07 [*95%-CI* -0.16, 0.02], *p* = 0.137), though statistically not significant, i.e., the modulating effect of CSF IL-6 on CSO-PVS rate of change did not differ significantly between those groups. Subsequent LME analysis with A-T- as reference group, confirmed a lower but non-significant interaction of time and CSF IL-6 levels in A-T-on CSO-PVS change over time (*B* = 0.05 [*95%-CI* -0.02, 0.12], *p* = 0.188, **Figure 4E; Supplementary table S7**). Similarly, we did not observe different interactive effects of CSF IL-6 on BG-PVS rate of change between A+T+ and A-T- (time x CSF IL-6 x A-T-: *B* = -0.03 [*95%-CI* -0.06, 0.01], *p* = 0.146), and subsequent LME analysis with A-T- as reference group revealed a marginal modulation of BG-PVS rate of change in dependence of CSF IL-6 levels in these subjects (time x CSF IL-6: *B* = 0.02 [*95%-CI* 0.00, 0.05], *p* = 0.078, **Figure 4F**; **Supplementary table S8**).The findings in A-T-align with the interaction effects of time by CSF IL-6 levels we found in the presumably healthy subsample (*see* 3.3.1.4 with **Figures 2E,F**)

**Figure 4.**
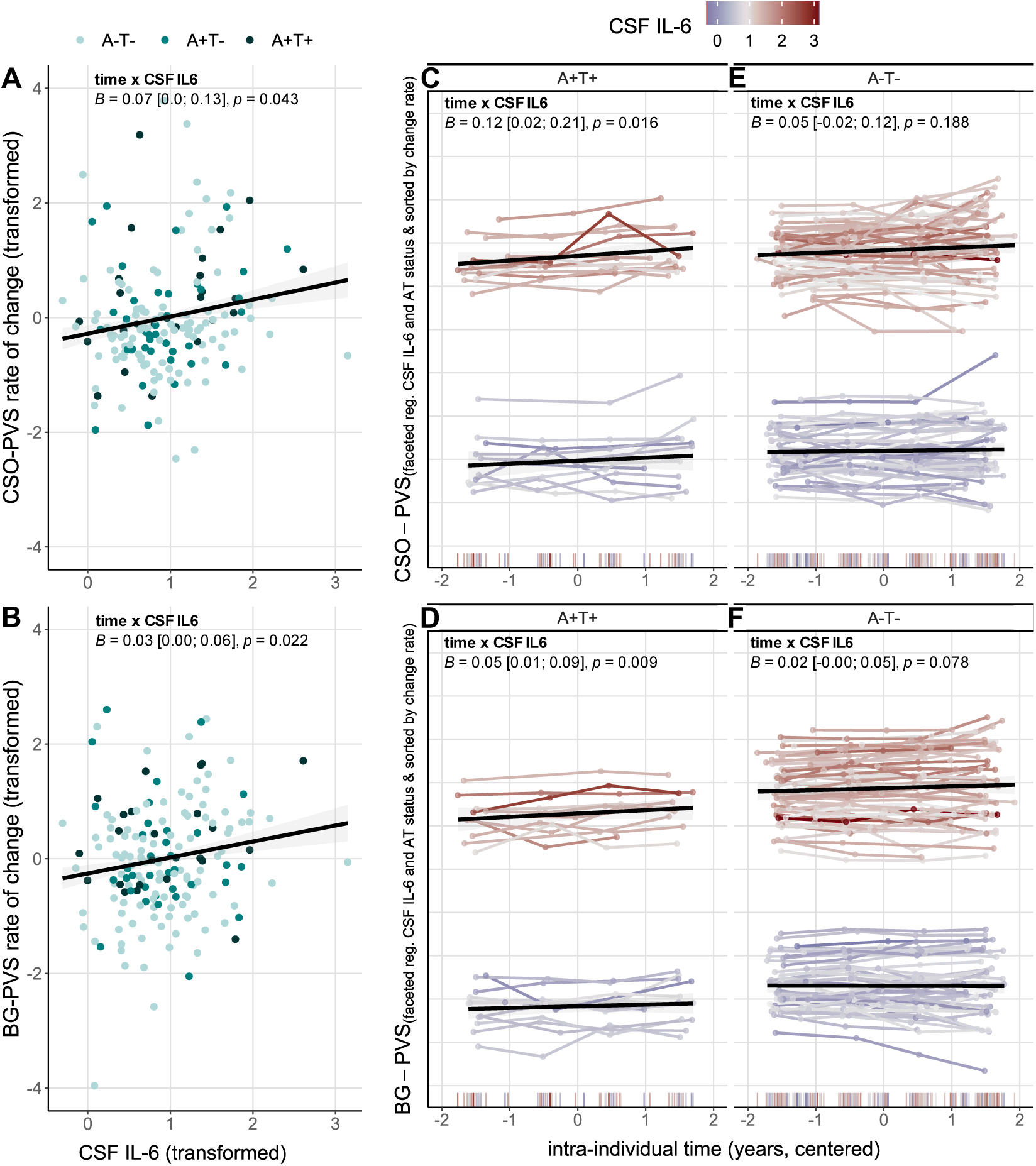
Association between CSF inflammatory marker Interleukin-6 and PVS rates of change and its dependency on AD biomarker profile. CSO-PVS in upper panel, BG-PVS in lower panel. (**A**-**B**) Higher levels of baseline IL-6 levels are associated with higher rates of change of both CSO- and BG-PVS in the whole sample. For easier visual comparison of this association across regions, rates of change of PVS volumes in CSO and BG, respectively, were z-transformed. CSF IL-6 levels were log-transformed. (**C-E**) Associations of CSF IL-6 and PVS rates of change are modulated by AD biomarker profile. (**C**) In A+T+, higher CSF IL-6 are related to higher rates of change of CSO-PVS. (**D**) In A+T+, higher CSF IL-6 are related to higher rates of change of BG-PVS. (**E**) In A-T-, CSO-PVS rates of change are not significantly modulated by CSF IL-6 levels. (**F**) Higher BG-PVS rates of change are marginally related to higher CSF IL-6 levels in A-T-subjects. Interindividual trajectories are shown. For readability, plots are faceted by AD biomarker profile and by low or high CSF IL-6 levels, based on median split on log-transformed CSF IL-6 levels (median = 0.85). Colours of individual trajectories correspond to CSF IL-6 levels. Cool colours depict lower, hot colours higher levels of CSF IL-6. We furthermore sorted individual trajectories by individual rate of change. Presented statistics are based on two-way interactions of time and CSF IL-6 levels of LME with respective AD biomarker profile as reference group.

##### 3.4.2.2 Associations between CSF IL-6 and AD biomarker profiles, hypertension, and WMH

CSF IL-6 levels did neither differ significantly across AD biomarker profiles (*Χ²*(2) = 0.256, *p* = 0.88, *η²* = 0.001) nor between treated hypertensive and normotensive individuals (*W* = 3706, *p* = 0.341, *r* = 0.071). We also found no indication of CSF IL-6 being associated with hippocampal volume at baseline (*ρ_spearman_* = 0.052, *p_uncorrected_* = 0.486) or with hippocampal rates of change (*ρ_spearman_* = -0.05, *p_uncorrected_* = 0.491). We did however observe IL-6 to relate to WMH, particularly for the later stages of the AD pathology, i.e. A+T+ group (CSO: *ρ_spearman_* = 0.23, *p_uncorrected_* = 0.25; BG: *ρ_spearman_* = 0.27, *p_uncorrected_* = 0.173), albeit not statistically significant (**Supplementary Figure S9**).

## 4 Discussion

We studied PVS enlargement over a three-year period in 557 subjects across syndromal cognitive stages of AD from a large German cohort to characterise longitudinal PVS dynamics and pinpoint potential exacerbators. We contributed to current efforts to elucidate vascular contributions to neurodegeneration from both methodological and medical standpoints(32). First, we demonstrated that studying PVS computationally and longitudinally in a large-scale multicentre study is technically feasible and reliable. Second, we provided longitudinal evidence of (a) PVS becoming more voluminous during ageing; (b) BG-PVS enlargement relating to hypertension; (c) CSO-PVS enlargement relating to AD diagnosis and ATN biomarkers; and (d) steeper PVS progression associated with an inflammatory biomarker and baseline WMH. The stability of longitudinal measurements as well as their meaningful associations with various factors imply that PVS could be a promising additional biomarker in studying and understanding healthy as well as pathological ageing.

### 4.1 PVS enlarge in the ageing process

In our large presumably healthy ageing cohort, CSO-PVS and BG-PVS volumes increased over follow-ups, consistent with previous studies(33–35), at an approximate rate of 7.55 and 17.11 mm³/year, respectively. BG-PVS enlargement was associated with higher age, although the extent of this enlargement was limited, as indicated by saturation effects seen in our data and previous work(3). The concurrent progression of white matter lesions or atrophy(6,60) might explain this finding. Sex and education were also tied to PVS enlargement. CSO-PVS volumes were greater in men, as described in previous cross-sectional studies(23,61). Despite the usage of fractional PVS volumes, this observation could be influenced by sex-dependent head size differences: individuals with larger heads require longer or thicker blood vessels to sustain adequate blood supply to the brain(62). Thus, alterations and injuries to their cerebral microvasculature can be expected to lead to higher PVS volumes. Moreover, participants with higher educational attainment had lower BG-PVS volumes. These prominent individual differences could be explained by engaging a healthier lifestyle(63). Lastly, we showed higher heterogeneity in CSO-PVS as compared to BG-PVS, indicating that CSO-PVS enlargement is influenced more strongly by individual differences.

### 4.2 PVS and hypertension

Cardiovascular risk factors can promote microvascular alterations and injuries(6). Indeed, participants of our presumably healthy subsample with a history of hypertension had higher rates of change of BG-PVS(18,36,64). This finding resonates, firstly, with the association of BG-PVS enlargement with WMH and, secondly, with the relation of lower BG-PVS to higher education and its presumed reflection of a healthier lifestyle (hypertensive individuals indeed had lower levels of education than their normotensive counterparts (*W*=43390, *p*=0.007, *r*=0.109)). Yet, the effect of hypertension on BG-PVS enlargement was subtle and not prominent in cross-sectional baseline measures. Study-specific factors might explain the absence of stronger associations: individuals with uncontrolled hypertension were excluded at recruitment and those with a history of hypertension were generally prescribed antihypertensive medication(65).

### 4.3 PVS and white matter hyperintensities

We observed that presumably healthy individuals with higher initial WMH volumes tend to exhibit greater PVS volumes, and that, over the subsequent years, WMH can form around PVS (see **Fig. 1E**), emphasising their interdependence and potentially shared underlying cardiovascular or ageing-related mechanisms(18,54,66). Additionally, baseline WMH and PVS rates of change of CSO and BG, respectively, correlated with one another, suggesting that future studies should explore the hypothesis of recurrent time-lagged associations. PVS have been in fact postulated as an early biomarker of cerebral small vessel alterations(33,35), which can precede WMH development(54) and accelerate white matter and grey matter deterioration (*for a review, see*(67)). We note nonetheless that focus on change-change modelling is required for further improving our understanding of the chronologic ties between PVS and WMH dynamics.

The fact that WMH spatially co-occur and can form around PVS poses a methodological challenge for assessments relying solely on T1w or T2w, as PVS and WMH are methodologically indistinguishable in these modalities(48) and, therefore, claims regarding PVS enlargement cannot be trusted since estimates will incorporate mixed effects(45). Though conservative, this obstacle stresses the need for multimodal quantification methodologies, such as that presented in this work, to reliably study PVS and their unique contributions to neurodegeneration and cognition.

### 4.4 PVS progression: influence of inflammation beyond Alzheimer’s pathology

CSO-PVS rates of change became steeper along syndromal cognitive stages and along the AD biomarker continuum, suggesting PVS enlargement may be a structural response to increased neurotoxic waste deposition(8). Conversely, BG-PVS progression appeared unrelated to AD, thereby lending credence to the ongoing discussion about the spatial heterogeneity of PVS aetiology(18,19,21,64), in which BG-PVS enlargement reflects ageing and cardiovascular risk, whereas CSO-PVS enlargement reflects pathological ageing in the context of Alzheimer’s disease. Hence, regional differences in PVS changes might help assess to what extent vascular or neurodegenerative processes operate.

AD biomarkers alone explained, however, just a small portion of the variance in PVS rates of change—as indicated by the small effects sizes (*η²*≤0.02) for differences across groups. Inflammation, on the other hand, provided additional explanation for the observed variability in PVS dynamics, as higher CSF IL-6 levels were associated with faster increases in fractional PVS volumes over time.

The relationship between CSF IL-6 and CSO-PVS rate of change is—to some extent— modulated by AD biomarker status. Compared to A-T-, CSO-PVS rates of change in A+T+ accelerated more strongly with increased baseline levels of CSF IL-6 levels. Similar relations were observed for BG-PVS rates of change. The AD biomarker dependent differences in BG-PVS enlargement due to CSF IL-6 were, however, less prominent, suggesting a more general effect of inflammation-driven enlargement. Even though not statistically significant, another study(27) reported similar findings: higher correlation between CSO-PVS and microglial activation in AD and MCI versus controls, while microglial activation and BG-PVS was less correlated in AD and MCI as compared to controls. These findings suggest there might be additional regional differences in the relation between inflammation and neurotoxic waste accumulation(1,26).

PVS dynamics are thus at the crossroads of hypertension, inflammation, and AD pathology and we here propose the notion that subjects along the AD continuum might face two related pathological processes that contribute to PVS enlargement (**Figure 5**). The first one could be a vicious cycle arising from Aβ and tau accumulation, which would hinder glymphatic clearance(13–15,19), leading to further stagnation of neurotoxic waste products, and PVS enlargement. The second one could be another vicious cycle driven by (chronic) neuroinflammation, whereby for example the accumulation of peripheral immune cells and cytokines in the PVS—e.g. triggered by blood-brain barrier dysfunction and neuroinflammation due to e.g. hypertension— would lead to impaired glymphatic clearance(2,24,56) and, ultimately, to PVS enlargement. Chronification of these vicious cycles might, in the long run, cause further damage to brain tissues, leading e.g., to WMH formation.

**Figure 5.**
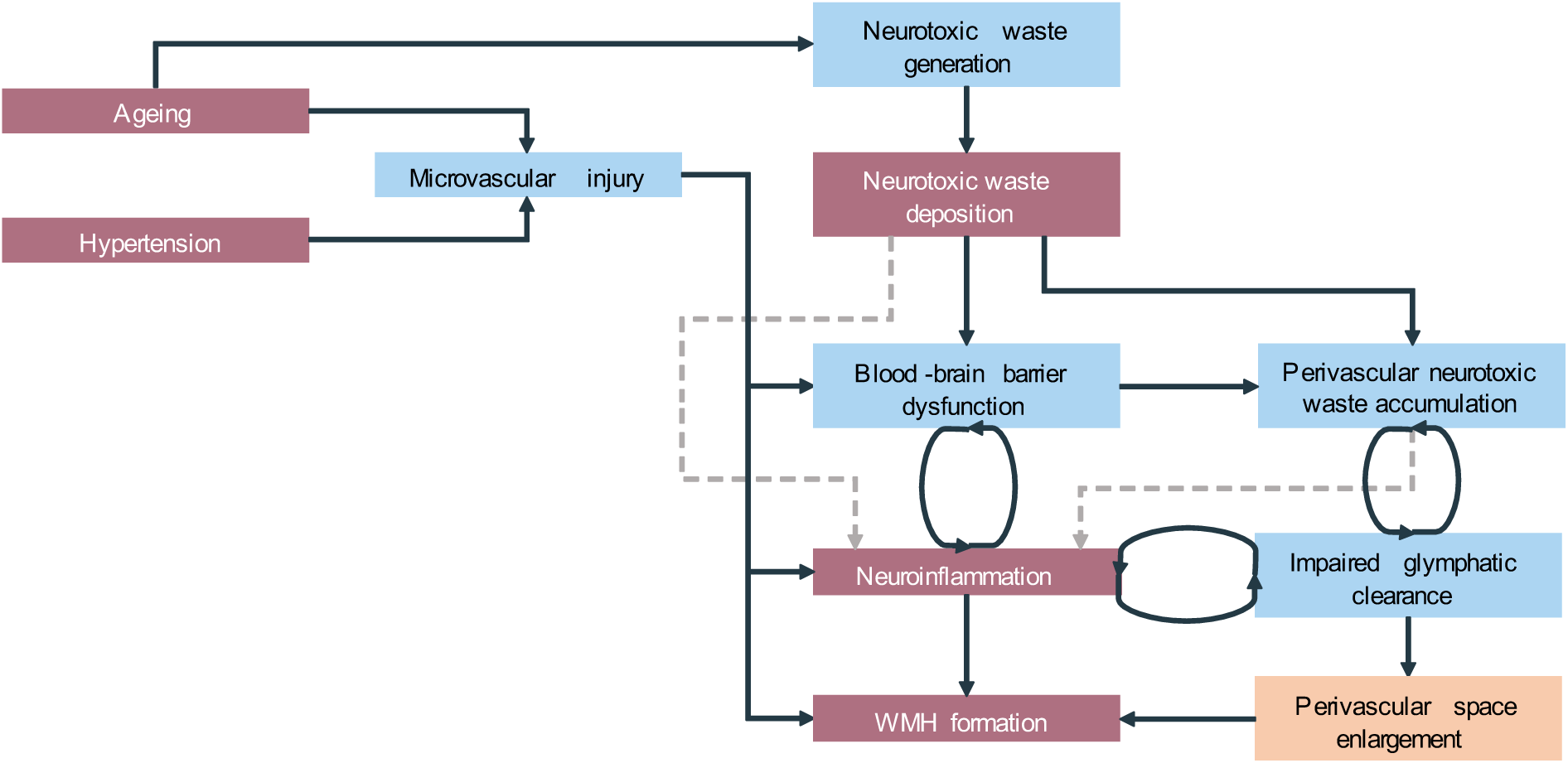
Proposed dual and interactive impact of AD pathology and inflammation on PVS enlargement. PVS enlargement might arise from two related vicious processes. On the one hand, neurotoxic waste (Aβ, tau) accumulation compromise glymphatic clearance, leading to drainage impairment and ultimately pathological enlargement of PVS. On the other hand, in ageing, hypertension, and AD, peripheral and neuroinflammation may interact to trigger blood-brain-barrier dysfunctions, which might cause accumulation of immune cells and cytokines in the PVS, likewise causing clearance impairments and pathological PVS enlargement. Chronification of these processes is thought to then lead to further damage to the brain tissue, culminating in the formation of white matter hyperintensities. Colour and line convention: light maroon and peach boxes indicate observed variables and those in light blue latent variables; dashed lines indicate relations proposed by literature, which we could not or only partly replicate in the present study. We note that this is a simplified, hypothetical model and additional variables and further (recurrent) relationships have not been considered. Moreover, for reasons of simplicity, we emphasise neurotoxic waste accumulation in terms of amyloid and tau. We do not rule out concurrent cascades driven by other neurotoxins. For reasons of readability, e.g., we here do not display possible recurrent relations between neurotoxic waste accumulation and neuroinflammation.

Taking into account previous findings(24,26,27,29), we consider conceivable that neuroinflammation might be indeed triggered in response to, prior, and during, accumulation of neurotoxic waste as well as the formation of amyloid plaques (dashed lines in **Figure 5**), despite our results only partly supporting this hypothesis. However, limited availability of CSF AD biomarkers and IL-6 reduced the power for this analysis considerably. Hence, we advise careful interpretation and encourage future modelling studies investigating the temporal cascade of these pathological processes, the enlargement of PVS, emergence of WMH, and their dynamic interrelations, as they would complement the the body of work establishing inflammatory processes as pivotal factors in AD beyond and in conjunction with amyloidosis, tauopathy, and neurodegeneration(24,26,29,68) and could provide helpful implications for risk assessment and targeting therapies.

### 4.5 Challenges and outlook

We identified four open questions. First, we noted that demographics explained a small portion of the variance in fractional PVS volumes, especially in CSO-PVS. Other cerebrovascular risk factors, lifestyle factors(69) or genetics(70) might contribute to explaining variability in baseline and longitudinal PVS measurements. Moreover, the exact underlying mechanisms for sex differences and particular explanations for the associations with education require further investigation. Second, while we assessed fractional volumes, additional morphological features can be investigated in future studies. This would enable determining whether PVS enlargement corresponds e.g., to a widening of the already-visible PVS or an increase in PVS counts. Both phenomena might be entangled and might have distinct clinical implications. Third, trends of increased PVS burden across syndromal cognitive stages suggest that PVS might serve as an early indicator for cognitive decline. Findings on that front are still conflicting(7,71) and require further careful investigation. Fourth, PVS dynamics do not seem disconnected from other neurodegenerative processes, making it sensible to examine their interactions over time to better understand disease pathogenesis and progression and identify clinical phenotypes that can help in deriving potential interventions.

## 5 Conclusion

Our findings contribute to the understanding of PVS progression as a biomarker in the context of neurodegenerative diseases. Ageing is a primary driver of PVS enlargement, but also detrimental cycles driven by cardiovascular risk, neurotoxic waste accumulation, and inflammation, contribute to PVS enlargement. Further research is needed to disentangle pathological cascades, their concurrent dynamics, and their unique contribution to disease progression. PVS might be an early marker of such pathophysiological cascades in AD and, through comprehensive understanding of these pathological dynamics, they have the potential to facilitate the development of early interventions.

## 6 Data availability statement

The data that support this study are not publically available, but may be provided upon reasonable request.

## 7 Acknowledgements

We would like to express our gratitude to all DELCODE participants. We also thank the Max Delbrück Centre for Molecular Medicine in the Helmholtz Association (MDC), Freie Universität Berlin Centre for Cognitive Neuroscience Berlin (CCNB), Bernstein Center für Computional Neuroscience Berlin, Universitätsmedizin Göttingen Core Facility MR-Research Göttingen, Institut für Klinische Radiologie Klinikum der Universität München, and Universitätsklinikum Tübingen MR-Forschungszentrum. We would like to thank Dr. Farshid Sepehrband for sharing his insights on PVS segmentation with us at the early stages of this project.

## 9 Author contributions

DELCODE study design: ED, AS, FJ

Conceptualisation: IM, JB, SS, GZ

Methodology: IM, JB, SS, GZ

Software: JB

Image processing: JB, RY

Image analysis: JB, PK, CA, MP, JG

Formal analysis: IM, JB

Investigation: IM, JB, SS, GZ

Writing – original draft preparation: IM, JB, SS, GZ

Writing – review and editing: all authors

