## Supplementary Material for "Perivascular space enlargement accelerates with hypertension, white matter hyperintensities, chronic inflammation, and Alzheimer’s disease pathology: evidence from a three-year longitudinal multicentre study"

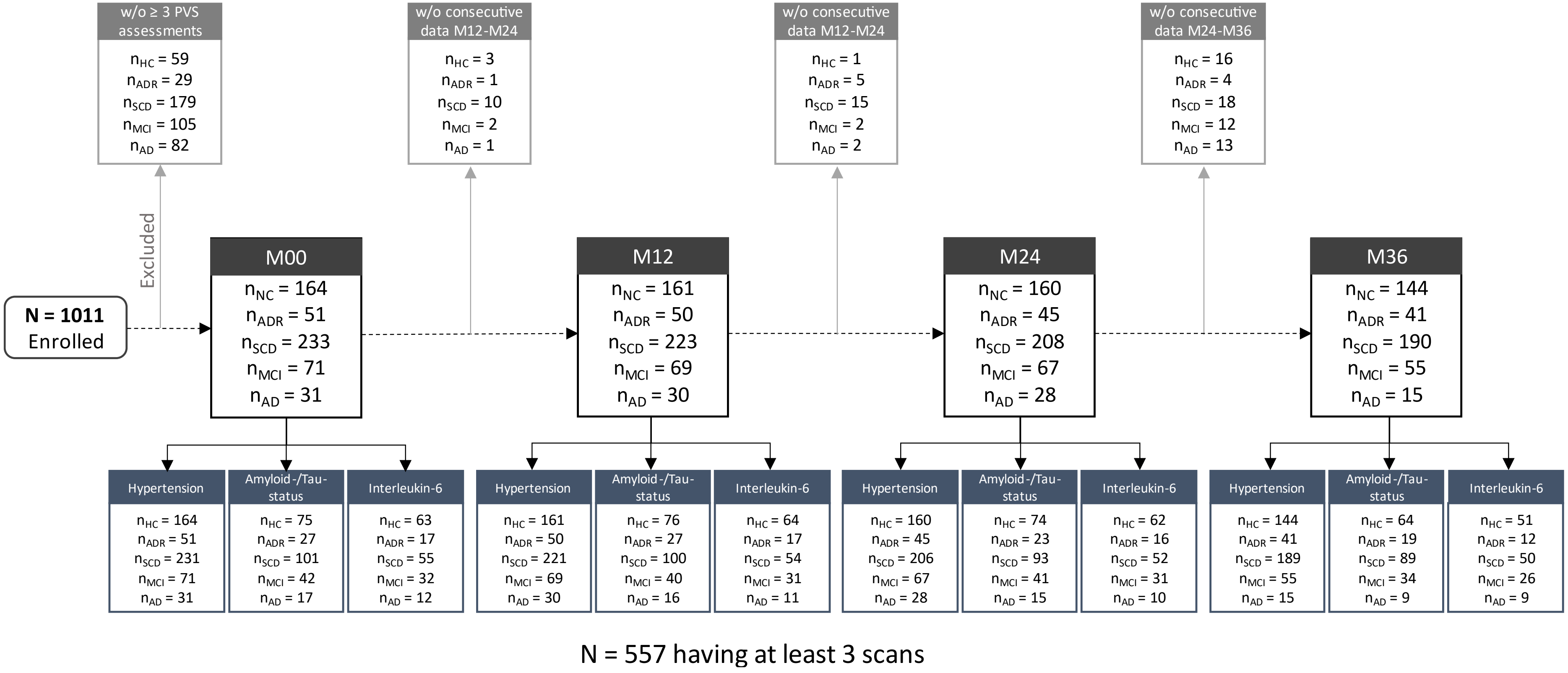


**Figure S1. Data availability flowchart across all four measurement time point**

### Supplementary information “computational PVS segmentation”

#### 1. Denoising

Many de-noising methods rely on the premise that the neighbourhood of a voxel contains sufficient information to recover the voxel’s original value—also referred to in the literature as the locally adaptive recovery paradigm[1] (e.g. mean and median filtering). The NLM filter, on the other hand, assumes images are redundant (there are regions within an image that are similar) and thus restores the intensity of a voxel based on non-adjacent yet similar regions. Let $f_0 : \Omega^3 \to \mathbb{R}$ and $f : \Omega^3 \to \mathbb{R}$ be the uncorrupted and corrupted versions of the same input scan and $\varepsilon \sim \mathcal{N}(0, \sigma^2)$ the additive white Gaussian noise corrupting the measurement, $f = f_0 + \varepsilon$. The NLM filter restores the intensity of a voxel $x_i \in \Omega^3$ via weighted averaging:

$$NL(f)(x_i) = \sum_{x_j \in \Omega^3} w(x_i, x_j) \cdot f(x_j), \qquad (1)$$

where $w(x_i, x_j)$ determines the weight of the voxel $x_j \in \Omega^3$ on $x_i$’s restoration. The weight of each voxel in the image is given by:

$$w(x_i, x_j) = \frac{1}{Z_i} e^{-\frac{\left\| f(N(x_i)) - f(N(x_j)) \right\|_2^2}{2 \cdot \beta \cdot \sigma^2 \cdot \#n(N_i)}}, \qquad (2)$$

where $Z_i \in \mathbb{R}$ is a normalising constant ensuring $\sum_{x_j} w(x_i, x_j) = 1$, $\beta \in \mathbb{R}$ a smoothing parameter, and $N(x)$ the neighbourhood around the voxel $x \in \Omega^3$, and $\#n(\cdot)$ the cardinality operator. Voxels with neighbourhoods comparable to that of the voxel of interest contribute more in the weighted average (Eq. 1).

In order to de-noise the input image adequately, a good estimate of the level of noise, given by $\sigma^2$, is necessary; otherwise blurring will be insufficient or excessive. Assuming redundancy, we can estimate this parameter directly from scans: if $N(x_i) \approx N(x_j)$ then $\left\| f_0(N(x_i)) + \varepsilon_{N(x_i)} - f_0\left(N(x_j)\right) \right\|_2^2 \approx 0$ and $\mathbb{E}\left[ \left\| f(N(x_i)) - f\left(N(x_j)\right) \right\|_2^2 \right] = \mathbb{E}\left[ \left\| f_0(N(x_i)) + \varepsilon_{N(x_i)} - f_0\left(N(x_j)\right) - \varepsilon_{N(x_j)} \right\|_2^2 \right] = 2\sigma^2$. Because the redundancy assumption does not always hold on clinical contexts[2], the local noise variance is calculated using a high-pass filtered version of the original image. To account for Rician noise typically observed in MRI, is rescaled based on image’s signal-to-noise ratio (see[2] for more details; code available at: https://sites.google.com/site/pierrickcoupe/softwares/denoising/mri-denoising?authuser=0).

#### 2.The Frangi filter: a multiscale Hessian-based filter

PVS segmentation often relies on Hessian analysis, which enables distinguishing between tubular structures and round or flat ones[3–5]. The operation of Hessian-based filters can be split into four major steps[5].

First, we convolve each input scan, $f: \Omega^3 \rightarrow \mathbb{R}$, with a family of Gaussian kernels, $\{\mathcal{G}_{\sigma_i^2} \mid \mathcal{G}_{\sigma_i^2}: \Omega^3 \rightarrow \mathbb{R} \wedge \sigma_i \in [\sigma_{min}, \sigma_{max}]\}$. Second, for each voxel and each kernel, we compute the Hessian matrix, $\left(H(f * \mathcal{G}_{\sigma_i^2})\right)_{ij} = \partial^2(f * \mathcal{G}_{\sigma_i^2}) / \partial x_i \partial x_j$. Third, we find the corresponding eigenvalues, $\lambda_{\sigma_i}^{(1)}, \lambda_{\sigma_i}^{(2)}, \lambda_{\sigma_i}^{(3)} \in \mathbb{R}$, with $|\lambda_{\sigma_i}^{(1)}| \leq |\lambda_{\sigma_i}^{(2)}| \leq |\lambda_{\sigma_i}^{(3)}|$. From here on, we exclusively examine regions with concave hypointense structures[5], i.e. where $\lambda_{\sigma_i}^{(2)}, \lambda_{\sigma_i}^{(3)} > 0$ in both T1w and FLAIR to rule out mistaking WMH for ePVS. Fourth, we compute the vesselness response function, which filters structures by sphericity, flatness, and contrast (**Figure S1**).

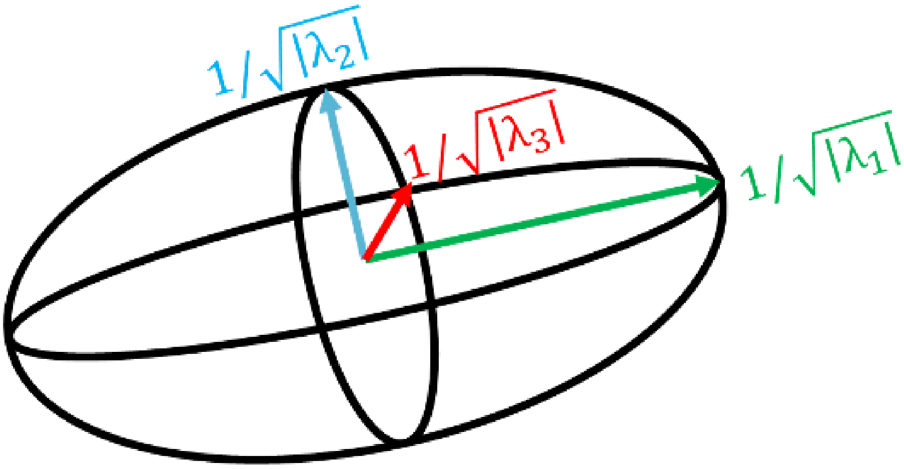


**Figure S2. Geometrical constraints that facilitate the search of PVS candidates.** To enhance tubular structures, Hessian-based filters capitalise on geometrical and visual properties encoded derived from the Hessian matrix, such as saliency, sphericity, and flatness. The Hessian matrix can be decomposed into its eigenvalues, $\boldsymbol{\lambda_1}, \boldsymbol{\lambda_2}, \boldsymbol{\lambda_3} \in \mathbb{R}$, with $|\boldsymbol{\lambda_1}| \leq |\boldsymbol{\lambda_2}| \leq |\boldsymbol{\lambda_3}|$ and eigenvectors, which geometrically describe the length and orientation of the axes of an ellipsoid, respectively. The search for PVS candidates is often limited to locations where $|\boldsymbol{\lambda_1}| \rightarrow \mathbf{0}$ and $|\boldsymbol{\lambda_1}|/\sqrt{|\boldsymbol{\lambda_2}\boldsymbol{\lambda_3}|} \rightarrow \mathbf{0}$ (i.e. containing elongated as opposed to spherical structures) and $|\boldsymbol{\lambda_2}|/|\boldsymbol{\lambda_3}| \rightarrow \mathbf{1}$ (i.e. containing linear as opposed to planar structures).

The Frangi filter[3]—a popular Hessian-based filtering method[5]—examines sphericity, $R_\beta = |\lambda_{\sigma_i}^{(1)}|/\sqrt{|\lambda_{\sigma_i}^{(2)}\lambda_{\sigma_i}^{(3)}|}$, flatness, $R_\alpha = |\lambda_{\sigma_i}^{(2)}|/|\lambda_{\sigma_i}^{(3)}|$, and contrast, $S = \sqrt{\lambda_{\sigma_i}^{(1)^2} + \lambda_{\sigma_i}^{(2)^2} + \lambda_{\sigma_i}^{(3)^2}}$ jointly in the so-called vesselness response function[3,5]:

$$V_{multiscale}(f; \alpha, \beta, \gamma, \sigma_{min}, \sigma_{max}) = \max_{\sigma \in [\sigma_{min}, \sigma_{max}]} \begin{cases} 0 \; if \lambda_{\sigma_i}^{(2)} < 0 \; or \lambda_{\sigma_i}^{(3)} < 0, \\ \left(1 - e^{-R_\alpha^2/2}\right) \cdot \left(e^{-R_\beta^2/2}\right) \cdot \left(1 - e^{-S^2/2\gamma^2}\right) \; otherwise, \end{cases} \quad (3)$$

where $\alpha$, $\beta$, and $\gamma \in \mathbb{R}^+$ determine the sensitivity of the filter to each of these geometrical and visual cues and $\sigma_{min}, \sigma_{max} \in \mathbb{R}^+$ relate to the size of the structures of interest (code available at: https://de.mathworks.com/matlabcentral/fileexchange/24409-hessian-based-frangi-vesselness-filter). We set up these five parameters to $\alpha = 0.5$, $\beta = 0.5$, and $\gamma$ to be half the value of the maximum Hessian norm, in line with the parameters suggested in the seminal work[3], and $\sigma_{min} = 0.1$ to $\sigma_{max} = 5$ to allow for the detection of structures of a wide range of sizes[6]—the distance between the zero-crossings of the Laplacian of the Gaussian kernel increases with $\sigma$, which permits enhancing larger objects of interest.

**3. Consensus map**

Though theoretically feasible, PVS segmentation using only T1w or T2w imaging is not ideal as WMH are likely to be mistakenly classified as PVS because their vesselness response exceeds that of PVS. Such classification errors bias subsequent analyses and interpretations, hindering determining the involvement of PVS alone on brain health function[7]. We, therefore, opted for a more conservative approximation in which we used both FLAIR and T1w imaging to better discern between these two neuroradiological features[8]: PVS appear concave and hypointense in both FLAIR and T1w images, whereas WMH appear convex and hyperintense in FLAIR and concave and hypointense in T1w images[8]. Mathematically speaking, we limited our search to voxels where $|\lambda_1| \approx 0$ and $\lambda_2 \approx \lambda_3 \gg 0$ in T1w and FLAIR. We kept the now-filtered T1w-based vesselness response as a reference for subsequent binarisation.

**4. Spatial processing for marginal modelling**

We registered all PVS segmentation maps to a DELCODE-specific Multi-Brain (MB) toolbox template[9] (code available at: https://github.com/WTCN-computational-anatomy-group/mb) and adjusted for local volume changes introduced by normalisation in PVS segmentation maps by modulation with Jacobian determinants[10,11]. PVS maps were smoothed with Gaussian kernels (6 mm full width at half maximum). Model was aligned with regional marginal models[12] (PVS ~ Time*AT profile + Age + Age² + Sex + Years of Education + Total Brain Volume).

**5. Quantification limitations**

This multimodal approximation to segment PVS and better discern them from WMH is conservative. We were able to demonstrate that our computational estimates resonated well with the clinical assessments (clinical visual ratings and manual counts; **Figure 1**).

Strikingly, we found a trend in the Bland-Altmann plots for PVS in the white matter: differences between computational and manual counts increased with more PVS. Such biases were also reported elsewhere[13,14] and are entirely methodologically driven: long tubular structures tend to be split into several segments, resulting in greater counts of PVS[7]. Although widespread, the accuracy of the Frangi filter can vary depending on the characteristics (vascular load) of the study population; other filters might yield better segmentation results in different scenarios[7]. Optimizing the automatic quantification of PVS hence still remains a matter for future research.

While useful, the multimodal method has drawbacks. First, its applicability depends on whether the sequences being jointly analysed have similar resolutions, given that differences in voxel size disallow visualising and quantifying PVS[7]. Second, sensitivity differences, co-registration errors as well as motion artefacts may compromise PVS identification, particularly when small[7,15]. Yet, the good stability of estimates we here achieved opposes such corruption. However, the selection of study-specific threshold values is nonetheless advisable. All in all, the development of automatic methods that can accurately identify different markers of cerebral small vessel alterations accurately requires more research. High-resolution multimodal techniques, as well as a reliable and comprehensive ground truth, may aid in reaching this goal. We emphasise that, for the time being, computerised segmentation offers a decent estimate of PVS burden, but its clinical assessment and validation remain essential.

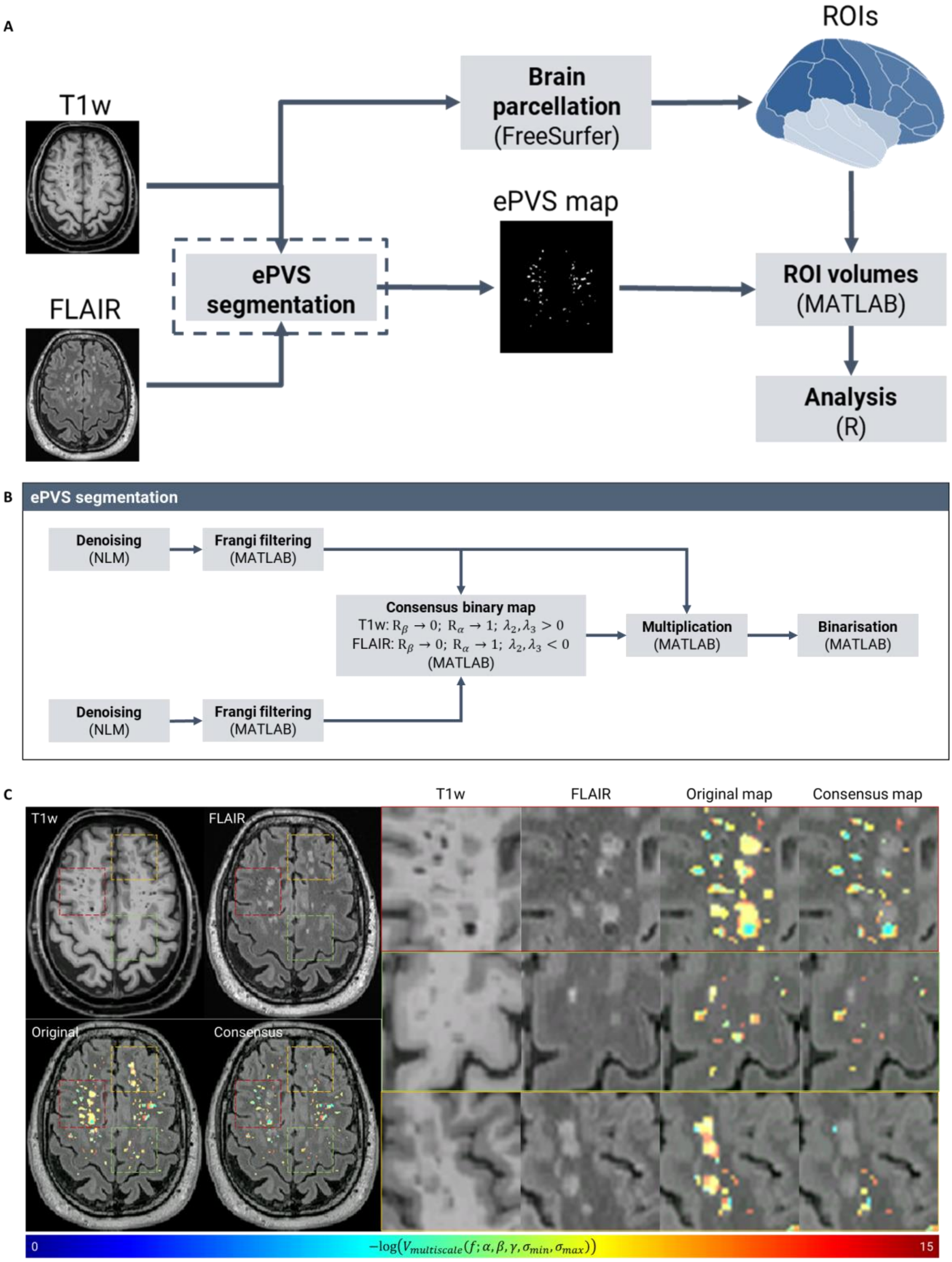

**Figure S3. PVS quantification and analysis pipeline.** (A) We segmented PVS based on baseline T1w and FLAIR scans. We then integrated PVS over basal ganglia and centrum semiovale and used R to study regional fractional PVS volume in relation to clinical variables and time. (B) Four key steps made up our PVS segmentation strategy. First, we used non-local means (NLM) to reduce the level of noise on T1w and FLAIR images. Second, we applied the Frangi filter to all images to enhance tubular structures. Third, to better distinguish between PVS and WMH, we fused the information provided by T1w and FLAIR imaging. Fourth, to finally produce PVS segmentation maps, we binarise the consensus maps. (C) Without our multimodality PVS segmentation approximation, (focal) WMH would be erroneously flagged as PVS if based solely on T1w scans (referred to as "Original map" in C).

**Table S4. Linear mixed effect modelling for temporal trajectories of hippocampal volume, showing different effects of time, of age, sex and years of education.** Models with correlated slope and random intercept: hippocampal volume ~ age + age² + time + sex + years of education + (1+ time | subject).

| | Hippocampal volume | | | |
|---|---|---|---|---|
| *Predictors* | *B* | *SE* | *CI* | *p* |
| (Intercept) | 0.20 | 0.06 | 0.09 – 0.31 | **<0.001** |
| age (linear) | -0.37 | 0.03 | -0.43 – -0.30 | **<0.001** |
| age (quadratic) | -0.04 | 0.03 | -0.11 – 0.02 | 0.160 |
| time | -0.08 | 0.00 | -0.08 – -0.07 | **<0.001** |
| sex | -0.43 | 0.07 | -0.56 – -0.30 | **<0.001** |
| years of education | 0.11 | 0.03 | 0.04 – 0.17 | **0.001** |
| $\sigma^2$ | 0.01 | | | |
| $\tau_{00}$ | 0.64 Subject | | | |
| $\tau_{11}$ | 0.00 Subject.time_ind | | | |
| $\rho_{01}$ | 0.54 Subject | | | |
| ICC | 0.99 | | | |
| N | 527 Subject | | | |
| Observations | 1927 | | | |
| Marginal $R^2$ / Conditional $R^2$ | 0.229 / 0.992 | | | |

Annotations. $\sigma^2$ = residual variance; $\tau_{00}$ = random intercept variance; $\tau_{11}$ = random slope variance; $\rho_{01}$ = covariance between random slope and intercept.

**Table S5. Linear mixed effect modelling for CSF IL-6 effects in presumably healthy subsample (CN and SCD).** Models with correlated slope and random intercept: PVS ~ time x CSF IL-6 + age + age² + sex + years of education (1+ time | subject).

Annotations. $\sigma^2$ = residual variance; $\tau_{00}$ = random intercept variance; $\tau_{11}$ = random slope variance; $\rho_{01}$ = covariance between random slope and intercept.

| | CSO-PVS | | | | BG-PVS | | | |
|---|---|---|---|---|---|---|---|---|
| *Predictors* | *B* | *SE* | *CI* | *p* | *B* | *SE* | *CI* | *p* |
| (Intercept) | -1.99 | 0.18 | -2.34 – -1.63 | <0.001 | -1.06 | 0.12 | -1.29 – -0.82 | **<0.001** |
| age (linear) | 0.03 | 0.08 | -0.13 – 0.19 | 0.706 | 0.12 | 0.05 | 0.01 – 0.23 | **0.028** |
| age (quadratic) | 0.07 | 0.09 | -0.10 – 0.23 | 0.439 | -0.05 | 0.06 | -0.16 – 0.06 | 0.382 |
| Years of education | 0.07 | 0.08 | -0.08 – 0.22 | 0.339 | -0.01 | 0.05 | -0.11 – 0.09 | 0.845 |
| sex | -0.24 | 0.15 | -0.54 – 0.05 | 0.108 | 0.16 | 0.10 | -0.03 – 0.36 | 0.099 |
| time | 0.02 | 0.05 | -0.07 – 0.11 | 0.712 | -0.01 | 0.02 | -0.05 – 0.02 | 0.448 |
| CSF IL-6 | -0.02 | 0.14 | -0.30 – 0.25 | 0.869 | 0.08 | 0.10 | -0.12 – 0.28 | 0.423 |
| Time x CSF IL-6 | 0.07 | 0.04 | -0.02 – 0.16 | 0.118 | 0.03 | 0.02 | -0.01 – 0.07 | 0.094 |
| **Random Effects** | | | | | | | | |
| $\sigma^2$ | 0.16 | | | | 0.02 | | | |
| $\tau_{00}$ | 0.50 Subject | | | | 0.24 Subject | | | |
| $\tau_{11}$ | 0.02 Subject.time_ind | | | | 0.00 Subject.time_ind | | | |
| $\rho_{01}$ | 0.25 Subject | | | | 0.40 Subject | | | |
| ICC | 0.76 | | | | 0.92 | | | |
| N | 114 Subject | | | | 113 Subject | | | |
| Observations | 422 | | | | 416 | | | |
| Marginal $R^2$ / Conditional $R^2$ | 0.052 / 0.773 | | | | 0.064 / 0.926 | | | |

**Table S6. Linear mixed effect modelling for CSO-PVS and BG-PVS in full sample, showing different trajectories over time, effects of age, sex and years of education.** Models with correlated slope and random intercept: PVS ~ age + age² + time + sex + years of education + (1+ time | subject).

| | CSO-PVS | | | | BG-PVS | | | |
|---|---|---|---|---|---|---|---|---|
| *Predictors* | *B* | *SE* | *CI* | *p* | *B* | *SE* | *CI* | *p* |
| (Intercept) | -1.72 | 0.06 | -1.83 – -1.60 | **<0.001** | -0.78 | 0.04 | -0.85 – -0.70 | **<0.001** |
| age (linear) | 0.07 | 0.04 | -0.00 – 0.15 | 0.052 | 0.17 | 0.02 | 0.13 – 0.22 | **<0.001** |
| age (quadratic) | -0.03 | 0.04 | -0.10 – 0.04 | 0.377 | -0.07 | 0.02 | -0.11 – -0.02 | **0.003** |
| time | 0.09 | 0.01 | 0.07 – 0.11 | **<0.001** | 0.04 | 0.00 | 0.03 – 0.05 | **<0.001** |
| Years of education | 0.02 | 0.04 | -0.05 – 0.09 | 0.629 | -0.05 | 0.02 | -0.10 – -0.01 | **0.018** |
| sex | -0.19 | 0.07 | -0.33 – -0.04 | **0.012** | -0.06 | 0.05 | -0.15 – 0.02 | 0.156 |
| **Random Effects** | | | | | | | | |
| $\sigma^2$ | 0.16 | | | | 0.03 | | | |
| $\tau_{00}$ | 0.60 Subject | | | | 0.24 Subject | | | |
| $\tau_{11}$ | 0.03 Subject.time_ind | | | | 0.00 Subject.time_ind | | | |
| $\rho_{01}$ | 0.24 Subject | | | | 0.22 Subject | | | |
| ICC | 0.79 | | | | 0.89 | | | |
| N | 522 Subject | | | | 531 Subject | | | |
| Observations | 1887 | | | | 1920 | | | |
| Marginal $R^2$ / Conditional $R^2$ | 0.033 / 0.801 | | | | 0.122 / 0.907 | | | |

Annotations. $\sigma^2$ = residual variance; $\tau_{00}$ = random intercept variance; $\tau_{11}$ = random slope variance; $\rho_{01}$ = covariance between random slope and intercept.

**Table S7. Linear mixed effect modelling for modulating effect of CSF IL-6 levels on CSO-PVS rates of change in dependence of biomarker profile.** Main model considered A+T+ as reference group (left). Subsequent analysis considered A-T- as reference group (right). Models with correlated slope and random intercept: CSO-PVS ~ age + age² + time + sex + years of education + AT + CSF IL-6 + CSF IL-6 x time x AT + (1+ time | subject).

| | **CSO-PVS** | | | | **CSO-PVS (A+T+ reference)** | | | | **CSO-PVS (A-T- reference)** | | | |
|---|---|---|---|---|---|---|---|---|---|---|---|---|
| *Predictors* | *B* | *SE* | *CI* | *p* | *B* | *SE* | *CI* | *p* | *B* | *SE* | *CI* | *p* |
| (Intercept) | -2.03 | 0.16 | -2.34 – -1.72 | **<0.001** | -1.61 | 0.21 | -2.03 – -1.19 | **<0.001** | -2.11 | 0.17 | -2.44 – -1.78 | **<0.001** |
| age (linear) | 0.12 | 0.07 | -0.02 – 0.26 | 0.095 | 0.08 | 0.07 | -0.06 – 0.23 | 0.269 | 0.08 | 0.07 | -0.06 – 0.23 | 0.269 |
| age (quadratic) | 0.02 | 0.07 | -0.13 – 0.16 | 0.830 | 0.03 | 0.07 | -0.12 – 0.17 | 0.720 | 0.03 | 0.07 | -0.12 – 0.17 | 0.720 |
| Years of education | 0.09 | 0.07 | -0.04 – 0.22 | 0.188 | 0.08 | 0.07 | -0.06 – 0.21 | 0.253 | 0.08 | 0.07 | -0.06 – 0.21 | 0.253 |
| time | -0.10 | 0.14 | -0.36 – 0.17 | 0.468 | -0.12 | 0.14 | -0.39 – 0.15 | 0.398 | -0.12 | 0.14 | -0.39 – 0.15 | 0.398 |
| sex | 0.02 | 0.04 | -0.05 – 0.09 | 0.585 | 0.02 | 0.04 | -0.05 – 0.09 | 0.533 | 0.02 | 0.04 | -0.05 – 0.09 | 0.533 |
| CSF IL-6 | 0.12 | 0.11 | -0.09 – 0.34 | 0.258 | 0.12 | 0.11 | -0.09 – 0.33 | 0.265 | 0.12 | 0.11 | -0.09 – 0.33 | 0.265 |
| time × CSF Il-6 | 0.07 | 0.03 | 0.00 – 0.13 | **0.043** | 0.12 | 0.05 | 0.02 – 0.21 | **0.016** | 0.05 | 0.04 | -0.02 – 0.12 | 0.188 |
| A-T- | | | | | -0.50 | 0.18 | -0.86 – -0.14 | **0.006** | | | | |
| A+T- | | | | | -0.49 | 0.21 | -0.90 – -0.08 | **0.019** | 0.01 | 0.16 | -0.31 – 0.32 | 0.970 |
| A+T+ | | | | | | | | | 0.50 | 0.18 | 0.14 – 0.86 | **0.006** |

| | | | | | | | | | | | | |
|---|---|---|---|---|---|---|---|---|---|---|---|---|
| time × CSF Il-6 x A-T- | | | | | -0.07 | 0.05 | -0.16 – 0.02 | 0.137 | | | | |
| time × CSF Il-6 x A+T- | | | | | -0.04 | 0.06 | -0.15 – 0.07 | 0.477 | 0.03 | 0.04 | -0.06 – 0.12 | 0.513 |
| time × CSF Il-6 x A+T+ | | | | | | | | | 0.07 | 0.05 | -0.02 – 0.16 | 0.137 |
| **Random Effects** | | | | | | | | | | | | |
| $\sigma^2$ | 0.17 | | | | 0.17 | | | | 0.17 | | | |
| $\tau_{00}$ | 0.61 Subject | | | | 0.58 Subject | | | | 0.58 Subject | | | |
| $\tau_{11}$ | 0.02 Subject.time_ind | | | | 0.02 Subject.time_ind | | | | 0.02 Subject.time_ind | | | |
| $\rho_{01}$ | 0.19 Subject | | | | 0.16 Subject | | | | 0.16 Subject | | | |
| ICC | 0.79 | | | | 0.79 | | | | 0.79 | | | |
| N | 168 Subject | | | | 168 Subject | | | | 168 Subject | | | |
| Observations | 617 | | | | 617 | | | | 617 | | | |
| Marginal $R^2$ / Conditional $R^2$ | 0.051 / 0.804 | | | | 0.088 / 0.804 | | | | 0.088 / 0.804 | | | |

Annotations. $\sigma^2$ = residual variance; $\tau_{00}$ = random intercept variance; $\tau_{11}$ = random slope variance; $\rho_{01}$ = covariance between random slope and intercept.

**Table S8. Linear mixed effect modelling for modulating effect of CSF IL-6 levels on BG-PVS rates of change in dependence of biomarker profile.** Main model considered A+T+ as reference group (left). Subsequent analysis considered A-T- as reference group (right). Models with correlated slope and random intercept: BG-PVS ~ age + age² + time + sex + years of education + AT + CSF IL-6 + CSF IL-6 x time x AT + (1+ time | subject).

| | **BG-PVS** | | | | **BG-PVS (A+T+ reference)** | | | | **BG-PVS (A-T- reference)** | | | |
|---|---|---|---|---|---|---|---|---|---|---|---|---|
| *Predictors* | *B* | *SE* | *CI* | *p* | *B* | *SE* | *CI* | *p* | *B* | *SE* | *CI* | *p* |
| (Intercept) | -0.91 | 0.09 | -1.09 – -0.72 | **<0.001** | -0.84 | 0.13 | -1.09 – -0.59 | **<0.001** | -0.84 | 0.10 | -1.03 – -0.64 | **<0.001** |
| age (linear) | 0.12 | 0.04 | 0.04 – 0.21 | **0.003** | 0.14 | 0.04 | 0.06 – 0.23 | **0.001** | 0.14 | 0.04 | 0.06 – 0.23 | **0.001** |
| age (quadratic) | -0.10 | 0.04 | -0.18 – -0.01 | **0.022** | -0.10 | 0.04 | -0.18 – -0.01 | **0.021** | -0.10 | 0.04 | -0.18 – -0.01 | **0.021** |
| Years of education | -0.02 | 0.04 | -0.09 – 0.06 | 0.693 | -0.03 | 0.04 | -0.11 – 0.05 | 0.501 | -0.03 | 0.04 | -0.11 – 0.05 | 0.501 |
| sex | 0.07 | 0.08 | -0.08 – 0.23 | 0.344 | 0.04 | 0.08 | -0.12 – 0.20 | 0.619 | 0.04 | 0.08 | -0.12 – 0.20 | 0.619 |
| time | -0.01 | 0.01 | -0.04 – 0.02 | 0.523 | -0.01 | 0.01 | -0.04 – 0.02 | 0.556 | -0.01 | 0.01 | -0.04 – 0.02 | 0.556 |
| CSF IL-6 | -0.00 | 0.07 | -0.13 – 0.13 | 0.958 | -0.01 | 0.07 | -0.14 – 0.12 | 0.867 | -0.01 | 0.07 | -0.14 – 0.12 | 0.867 |
| time × CSF Il-6 | 0.03 | 0.01 | 0.00 – 0.06 | **0.022** | 0.05 | 0.02 | 0.01 – 0.09 | **0.009** | 0.02 | 0.01 | -0.00 – 0.05 | 0.078 |
| A-T- | | | | | 0.00 | 0.11 | -0.22 – 0.22 | 0.989 | | | | |
| A+T- | | | | | -0.19 | 0.13 | -0.44 – 0.06 | 0.137 | -0.19 | 0.10 | -0.38 – 0.00 | 0.050 |
| A+T+ | | | | | | | | | -0.00 | 0.11 | -0.22 – 0.22 | 0.989 |

| | | | | | | | | | | | | |
|---|---|---|---|---|---|---|---|---|---|---|---|---|
| time × CSF Il-6× A-T- | | | | | -0.03 | 0.02 | -0.06 – 0.01 | 0.146 | | | | |
| time × CSF Il-6 × A+T- | | | | | -0.02 | 0.02 | -0.07 – 0.02 | 0.324 | 0.00 | 0.02 | -0.03 – 0.04 | 0.809 |
| time × CSF Il-6 × A+T+ | | | | | | | | | 0.03 | 0.02 | -0.01 – 0.06 | 0.146 |
| **Random Effects** | | | | | | | | | | | | |
| $\sigma^2$ | 0.03 | | | | 0.03 | | | | 0.03 | | | |
| $\tau_{00}$ | 0.23 Subject | | | | 0.22 Subject | | | | 0.22 Subject | | | |
| $\tau_{11}$ | 0.00 Subject.time_ind | | | | 0.00 Subject.time_ind | | | | 0.00 Subject.time_ind | | | |
| $\rho_{01}$ | 0.37 Subject | | | | 0.38 Subject | | | | 0.38 Subject | | | |
| ICC | 0.90 | | | | 0.90 | | | | 0.90 | | | |
| N | 167 Subject | | | | 167 Subject | | | | 167 Subject | | | |
| Observations | 614 | | | | 614 | | | | 614 | | | |
| Marginal $R^2$ / Conditional $R^2$ | 0.081 / 0.907 | | | | 0.103 / 0.908 | | | | 0.103 / 0.908 | | | |

Annotations. $\sigma^2$ = residual variance; $\tau_{00}$ = random intercept variance; $\tau_{11}$ = random slope variance; $\rho_{01}$ = covariance between random slope and intercept.

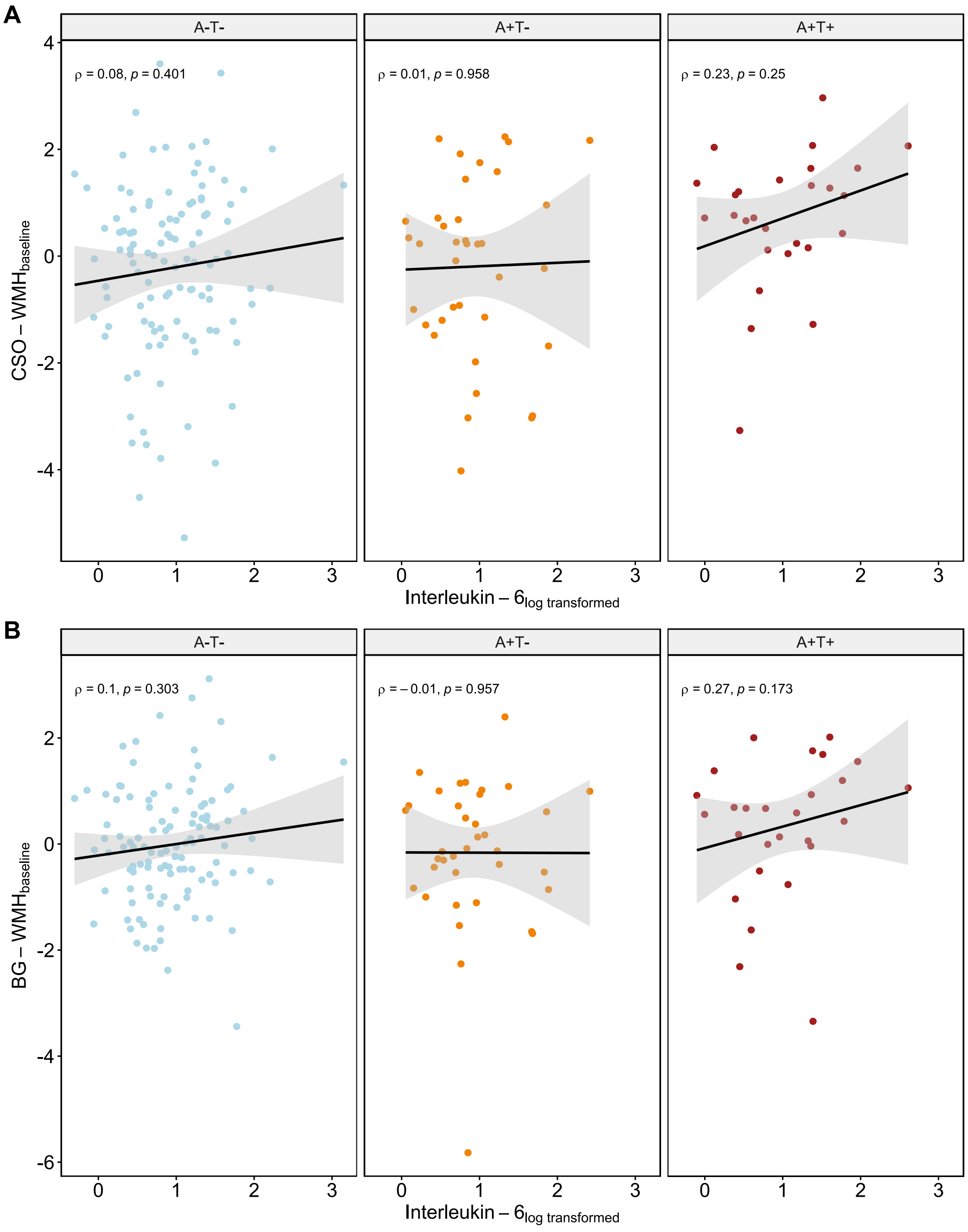


**Figure S9. Relations between CSF IL-6 levels and WMH at baseline**. Spearman correlations are shown for WMH in (A) CSO and (B) BG. WMH volumes were corrected for linear and quadratic age effects, sex and years of education.
